# Subtype-Specific Genetic Liability to Congenital Heart Disease and Late Atrial Fibrillation: A Cross-Cohort Mendelian Randomization Study with Quantification of Same-Cohort Bias

**DOI:** 10.64898/2026.11.25.26369016

**Authors:** Shuyu Wen, Fei Li, Yin Wang, Xingjian Hu

## Abstract

**Background.:** Atrial fibrillation (AF) is a common late complication after congenital heart disease (CHD) repair. Whether genetically determined CHD subtypes confer causal AF predisposition is unresolved. MR studies of rare congenital exposures often draw exposure and outcome from one biobank, biasing estimates toward observational associations. We performed a cross-cohort, subtype-resolved MR study and quantified this same-cohort bias.

**Methods.:** Genome-wide significant instruments (P<5×10⁻⁸; F>10; r²<0.001) for four CHD subtypes (BAV, LVOTO/CoA, ASD, overall CHD) came from FinnGen R13. Cross-cohort analyses paired these with independent GWAS Catalog outcomes: thoracic aneurysm (GCST90027266), AF (GCST006414), and heart failure (GCST009541). Inverse-variance weighted MR was primary; MR-Egger, weighted median, Steiger, leave-one-out, and MR-PRESSO were sensitivity analyses (Bonferroni across 12 tests). Parallel within-FinnGen analyses served only as a bias calibrator.

**Results.:** BAV and LVOTO/CoA instruments yielded modest, Bonferroni-robust AF effects (BAV OR 1.046, 95% CI 1.031–1.062, Bonferroni p=2.1×10⁻⁸; LVOTO OR 1.065, 1.042–1.088, p=1.0×10⁻⁷), concordant across sensitivity analyses; heart-failure effects were non-robust (Bonferroni p=0.259 and 0.266). Same-cohort aortopathy effects (BAV→aneurysm OR 1.45; LVOTO OR 1.77) were not reproduced in independent data (OR 0.935 and 0.889; confidence intervals excluding the same-cohort estimates), a gap exceeding eight standard errors. A negative 1p22 colocalization (six pairs, PP4<0.003) is reported separately.

**Conclusions.:** Aortopathy-prone CHD subtypes show modest, subtype-specific, Bonferroni-robust increases in late AF odds (odds ratios, not absolute risks). Same-cohort MR overstated aortopathy effects several-fold; cross-cohort replication should be standard in rare-disease MR. These findings support subtype-aware arrhythmia surveillance in BAV and CoA survivors.

**What Is New?:**

- In independent, non-overlapping cohorts, genetically determined BAV and LVOTO/CoA increase AF odds by approximately 5% and 7% (odds ratios, not absolute risk increases; Bonferroni p=2.1×10⁻⁸ and 1.0×10⁻⁷), with smaller, non-robust heart-failure effects.
- Large same-cohort aortopathy effects collapse to point estimates below unity in independent data, directly quantifying how far same-cohort MR can inflate effects for rare congenital exposures.

**What Are the Clinical Implications?:**

- Subtype-aware atrial-fibrillation surveillance may be warranted in BAV and CoA survivors, and cross-cohort replication should be required before single-cohort MR findings inform clinical practice.

## 1. Introduction

Congenital heart disease (CHD) affects approximately 1% of live births and is the most common class of birth defect.^[1]^ Advances in paediatric and congenital cardiac surgery have transformed CHD into a chronic condition, with most patients now surviving into adulthood.^[2]^ This demographic shift has unveiled a second epidemic—late cardiovascular morbidity among CHD survivors—of which atrial fibrillation (AF) is among the most common and clinically consequential.^[3,4]^

AF after CHD repair is associated with stroke, heart failure, and reduced survival, and its management is a growing burden in adult congenital heart disease (ACHD) clinics.^[3]^ Observational associations between CHD subtype and late AF are vulnerable to confounding by surgical era, residual lesions, and shared developmental pathology, as well as to reverse causation. Mendelian randomization (MR) uses germline genetic variants as instrumental variables to estimate causal effects while limiting these biases.^[7]^ Applying MR to CHD has to date been dominated by studies of maternal exposures as causes of CHD, or by analyses of CHD at the level of broad categories: a recent MR reported that congenital great-vessel anomalies increase AF risk (OR ≈ 1.07, 95% CI 1.02–1.12) and examined stroke outcomes.^[8]^ Whether genetically determined CHD subtypes differ in their causal predisposition to AF has not been resolved at the subtype level.

A second, methodological problem is specific to rare-disease MR. Because genome-wide significant instruments for individual CHD subtypes are available mainly from large biobank GWAS, such studies commonly take both exposure and outcome from the same cohort. Sample overlap in MR is known to pull estimates toward the corresponding observational association,^[16]^ so same-cohort estimates for rare congenital exposures may be systematically inflated. This issue is rarely quantified directly.

We therefore conducted a cross-cohort, subtype-resolved two-sample MR study to estimate the causal effects of genetically determined CHD subtypes on late AF using independent, non-overlapping outcome GWAS. We prespecified a comparison with a parallel within-FinnGen analysis to quantify same-cohort bias. Our primary hypothesis was that aortopathy-prone subtypes (BAV and LVOTO including CoA) carry a subtype-specific predisposition to AF; our secondary objective was to determine how much same-cohort design inflates subtype-specific MR estimates. The conceptual reverse direction—genetically determined aortic diameter increasing risk of LVOT (including CoA) CHD—was previously established in Circulation: Genomic and Precision Medicine,^[19]^ providing paradigm-level precedent for MR between aortic and CHD phenotypes.

## 2. Methods

### 2.1 Study design and data sources

This is a two-sample MR study conducted in accordance with STROBE-MR guidelines.^[9]^ Analyses were performed in R (v4.6.2) using TwoSampleMR (v0.7.14) for harmonisation and MR estimation, MRPRESSO (v1.0) for outlier detection, and coloc (v5.2.3) for colocalization. Because all analyses used only publicly available, de-identified summary-level data, institutional review board approval and individual informed consent were not required (exempt). The study adheres to the Transparency and Openness Promotion (TOP) guidelines: summary statistics are publicly available (FinnGen, https://www.finngen.fi; GWAS Catalog, https://www.ebi.ac.uk/gwas), analysis code is provided as supplementary material, and no individual-level data were accessed. The cross-cohort primary analysis pairs FinnGen exposures with independent GWAS Catalog outcomes to avoid sample overlap; the within-FinnGen analysis is presented only to quantify overlap bias.

Exposures — CHD subtypes (FinnGen R13). Summary statistics were obtained from FinnGen R13 (free public release; direct download from FinnGen Google Cloud Storage). Tetralogy of Fallot (ToF) was excluded from the primary analysis because it contained no genome-wide significant instruments (see 2.2 and Limitations). Remaining endpoints and their verified genome-wide significant instrument counts are listed in Table 1.

**Table 1.**
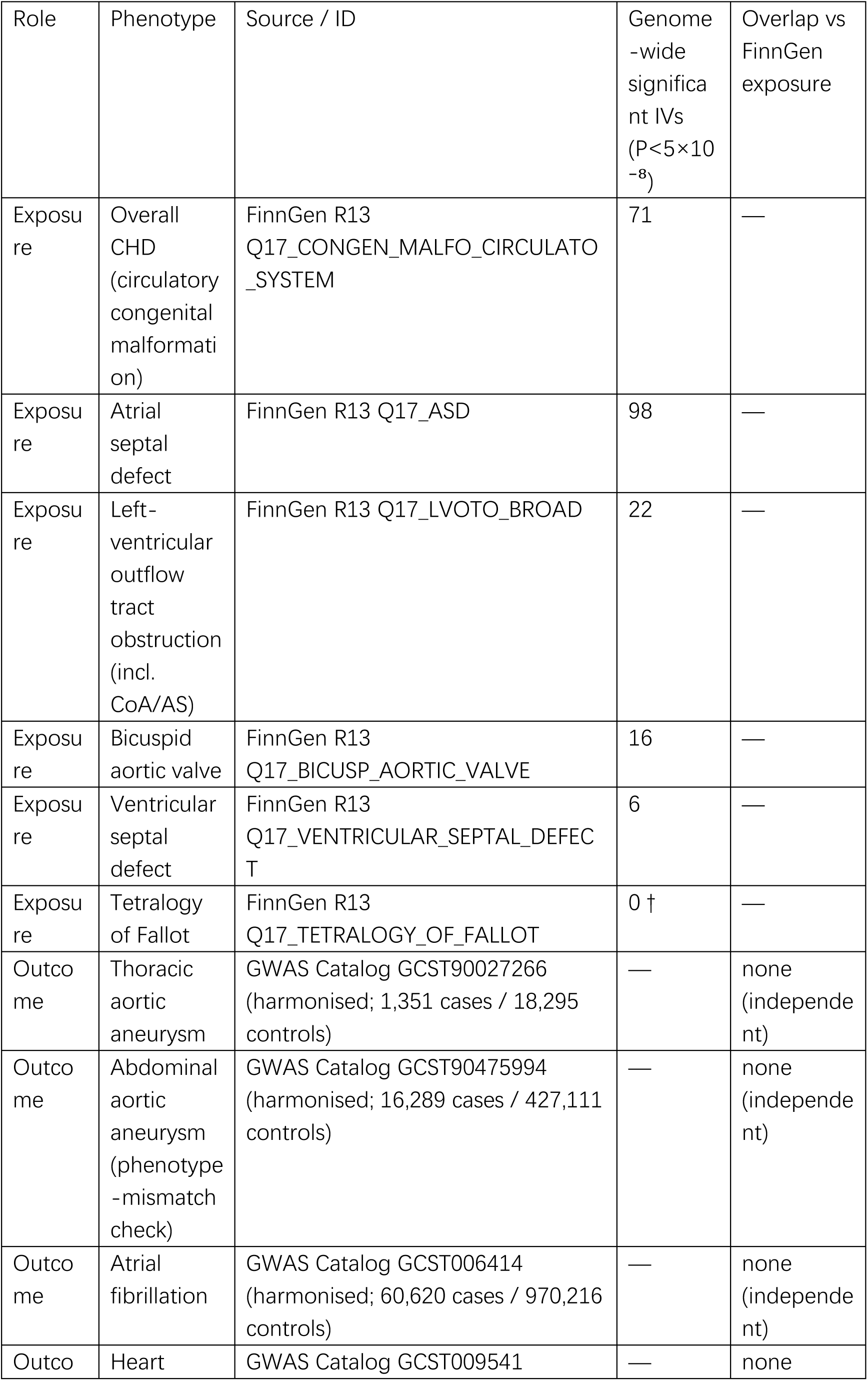
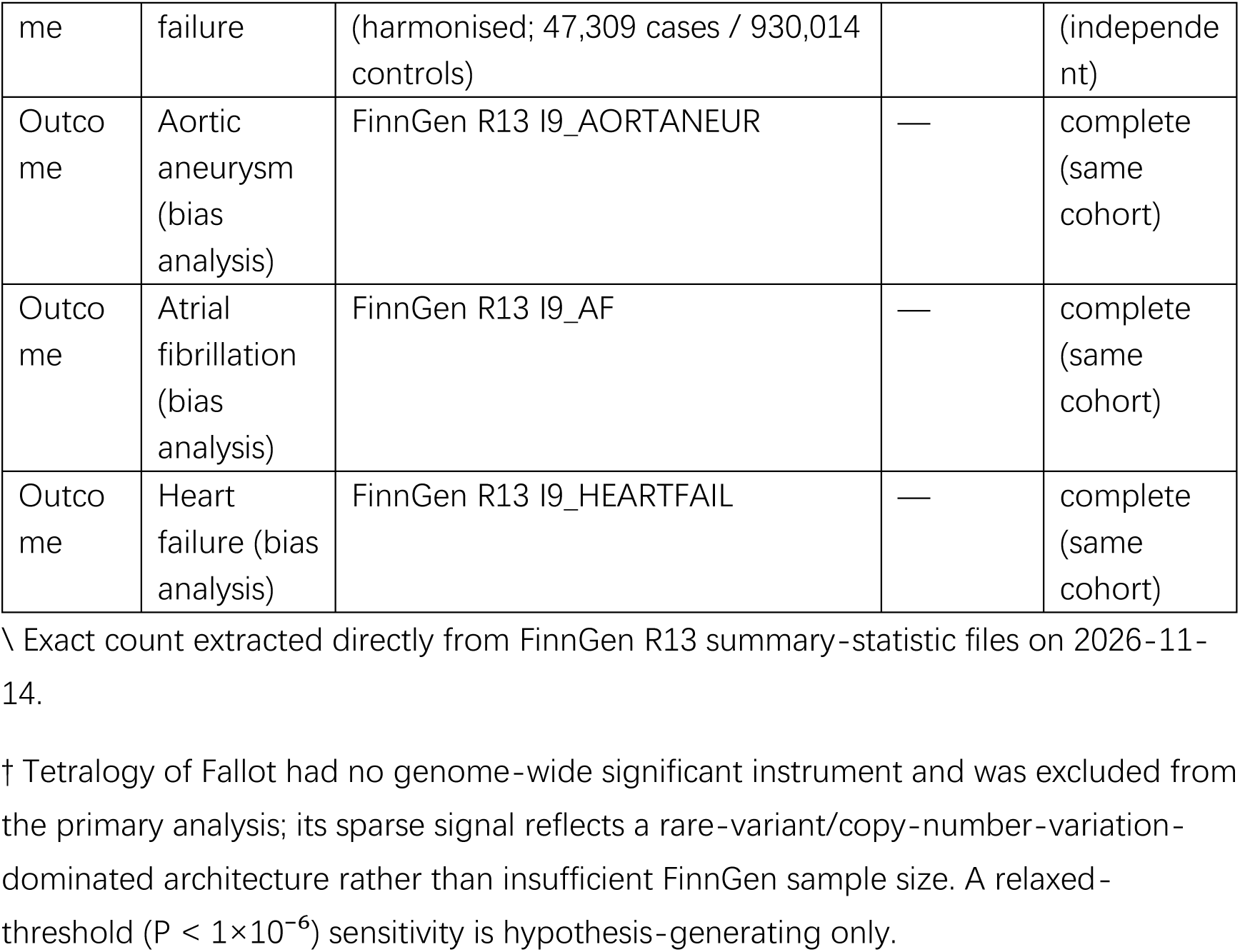
Data sources and verified instrument characteristics.

| Role | Phenotype | Source / ID | Genome-wide significant IVs (P<5×10<sup>-8</sup>) | Overlap vs FinnGen exposure |
|---|---|---|---|---|
| Exposure | Overall CHD (circulatory congenital malformation) | FinnGen R13 Q17_CONGEN_MALFO_CIRCULATO_SYSTEM | 71 | — |
| Exposure | Atrial septal defect | FinnGen R13 Q17_ASD | 98 | — |
| Exposure | Left-ventricular outflow tract obstruction (incl. CoA/AS) | FinnGen R13 Q17_LVOTO_BROAD | 22 | — |
| Exposure | Bicuspid aortic valve | FinnGen R13 Q17_BICUSP_AORTIC_VALVE | 16 | — |
| Exposure | Ventricular septal defect | FinnGen R13 Q17_VENTRICULAR_SEPTAL_DEFECT | 6 | — |
| Exposure | Tetralogy of Fallot | FinnGen R13 Q17_TETRALOGY_OF_FALLOT | 0 † | — |
| Outcome | Thoracic aortic aneurysm | GWAS Catalog GCST90027266 (harmonised; 1,351 cases / 18,295 controls) | — | none (independent) |
| Outcome | Abdominal aortic aneurysm (phenotype-mismatch check) | GWAS Catalog GCST90475994 (harmonised; 16,289 cases / 427,111 controls) | — | none (independent) |
| Outcome | Atrial fibrillation | GWAS Catalog GCST006414 (harmonised; 60,620 cases / 970,216 controls) | — | none (independent) |
| Outcome | Heart failure | GWAS Catalog GCST009541 (harmonised; 47,309 cases / 930,014 controls) | — | none (independent) |
| Outcome | Aortic aneurysm (bias analysis) | FinnGen R13 I9_AORTANEUR | — | complete (same cohort) |
| Outcome | Atrial fibrillation (bias analysis) | FinnGen R13 I9_AF | — | complete (same cohort) |
| Outcome | Heart failure (bias analysis) | FinnGen R13 I9_HEARTFAIL | — | complete (same cohort) |

\ Exact count extracted directly from FinnGen R13 summary-statistic files on 2026-11-14.
† Tetralogy of Fallot had no genome-wide significant instrument and was excluded from the primary analysis; its sparse signal reflects a rare-variant/copy-number-variation-dominated architecture rather than insufficient FinnGen sample size. A relaxed-threshold (P < 1×10<sup>-6</sup>) sensitivity is hypothesis-generating only.

Outcomes — late cardiovascular complications (independent cohorts). Cross-cohort primary outcomes were GWAS Catalog harmonised summary statistics from independent consortia: thoracic aortic aneurysm (GCST90027266; 1,351 cases / 18,295 controls, as reported in the GWAS Catalog discovery-sample description for Roychowdhury et al., Am J Hum Genet 2021, PMID 34265237), AF (GCST006414; 60,620 cases / 970,216 controls), and HF (GCST009541; 47,309 cases / 930,014 controls). To test the phenotype-mismatch explanation for aortopathy non-replication, an additional independent harmonised abdominal aortic aneurysm GWAS (GCST90475994; 16,289 cases / 427,111 controls) was analysed as a prespecified check; because its standard_error field was entirely missing, standard errors for log(OR) were recovered from the reported 95% confidence intervals as SE = [ln(CI_upper) − ln(CI_lower)]/3.92 (Supplementary Table S6). For the within-cohort bias analysis, FinnGen R13 endpoints (aortic aneurysm I9_AORTANEUR, AF I9_AF, HF I9_HEARTFAIL, VTE I9_VTE) were used.

Outcome definitions and the age window. Each FinnGen outcome is defined by a registry-and inpatient-based ICD code set (e.g., AF: ICD-10 I48; HF: I50; aortic aneurysm: I71 excluding I71.0). A caveat applies to the late-complication framing: CHD exposures are predominantly paediatric in onset, whereas AF, HF, and aneurysm typically manifest in adulthood. If an endpoint is defined without an age lower bound, the large number of young, event-free exposed individuals contributes to the control arm and can bias MR estimates toward the null (follow-up-incompleteness bias). FinnGen has precedent for this issue—for example, the endpoint “Aortic valve malfunction, age under 60 years” (I9_AORTVALV_EARLY) applies an EVENT_AGE<60 precondition to separate early-onset from degenerative disease^[18]^—and the relevant age windows must be verified in the FinnGen definition repository. Where no age restriction is applied, estimates are interpreted as the effect of genetically determined CHD liability on lifetime cumulative complication risk, not a time-window-specific absolute risk.

### 2.2 Instrument selection

SNPs associated with each CHD subtype at genome-wide significance (P < 5×10⁻⁸) were selected as instruments. Linkage disequilibrium clumping was performed (r² < 0.001 within 10 Mb) using the 1000 Genomes European reference panel. Variants with minor allele frequency < 1% were excluded. Instrument strength was evaluated by the F-statistic (F = β²/se²); instruments with F > 10 were retained.^[10]^ ToF yielded zero genome-wide significant variants and was excluded. VSD (6 instruments) was retained but flagged as weak-evidence. Harmonisation removed palindromic SNPs with ambiguous strand orientation and intermediate allele frequency; specifically, following the TwoSampleMR (v0.7.14) default (action=2, tolerance=0.08), palindromic variants with effect-allele frequency between 0.42 and 0.58 were excluded.

### 2.3 MR estimation

The validity of an MR estimate rests on three core instrumental-variable assumptions: (i) relevance—the variants are robustly associated with the exposure (all retained instruments had F>10; §2.2); (ii) independence—the variants are independent of confounders of the exposure–outcome association; and (iii) exclusion restriction—the variants affect the outcome only through the exposure. The primary estimate used inverse-variance weighted (IVW) MR, which assumes all instruments satisfy the exclusion restriction.^[11]^ Sensitivity analyses were MR-Egger regression (pleiotropy-robust, with intercept test), weighted median (consistent if ≥50% of weight is from valid instruments), and weighted mode.^[12,13]^ Estimates are reported as odds ratios (OR) per genetically determined CHD subtype with 95% confidence intervals.

### 2.4 Sensitivity and robustness

Heterogeneity was assessed with Cochran’s Q, horizontal pleiotropy with the MR-Egger intercept and MR-PRESSO outlier test,^[14]^ and influence with leave-one-out analysis (full results across all methods are provided in Supplementary Table S1; MR-PRESSO outputs in Supplementary Table S2; leave-one-out analyses in Supplementary Table S3). Steiger directionality testing^[15]^ was performed using phenotype-specific case/control counts from FinnGen R13 (BAV 1,356; LVOTO 4,050; ASD 1,443; overall CHD 6,216; total N=500,186) and outcome GWAS counts; because all traits were binary, SNP–trait correlations were computed on the liability scale with get_r_from_lor() rather than the quantitative-trait default (Supplementary Table S4). A Bonferroni-corrected threshold was applied across the 12 primary tests.

### 2.5 Quantifying same-cohort bias

Because the primary design is cross-cohort and free of sample overlap, the within-FinnGen analysis (exposures and outcomes both from FinnGen R13) was run in parallel solely to quantify overlap bias. Adjudication rule: the cross-cohort estimate is the primary result. Where the within-FinnGen estimate is larger in magnitude, the difference is interpreted as overlap bias (sample overlap, winner’s curse, and shared controls can inflate MR estimates toward observational associations^[16]^), not as a stronger causal signal. The direction and magnitude of this inflation are reported explicitly rather than treated as a nuisance. The analysis was not formally preregistered; however, the overall analysis plan—subtype-resolved two-sample MR with IVW as the primary estimator, the sensitivity analyses described above, Bonferroni correction across 12 primary tests, and an aortopathy-focused hypothesis for BAV and LVOTO—was documented in an internal study plan dated before data extraction (14 November 2026; available from the corresponding author on reasonable request). The cross-cohort design was adopted from the outset specifically to avoid sample overlap, and the within-cohort comparison is therefore assigned the explicit adjudication rule stated above rather than being treated as an alternative causal analysis.

### 2.6 Colocalization of the 1p22 instrument locus

To assess whether the 1p22 instruments act through PALMD, we performed Bayesian colocalization with coloc (v5.2.3; default priors p1=p2=1×10⁻⁴, p12=1×10⁻⁵). Because nearly all BAV/LVOTO lead instruments map to this single locus (Table S5), we extracted all variants in chr1:99.0–100.2 Mb (GRCh38) from the FinnGen R13 BAV and LVOTO summary statistics and tested each against PALMD (ENSG00000099260) cis-eQTLs from three GTEx tissues in the eQTL Catalogue (aorta QTD000131, atrial appendage QTD000251, left ventricle QTD000256). Effect alleles were harmonised across datasets and palindromic/ambiguous variants removed, leaving ∼3,850 shared SNPs per pair. We report the posterior probabilities of the five coloc hypotheses (H0–H4), with PP4 ≥ 0.8 taken as support for a shared causal variant. As the harmonised outcome files lacked effect-allele frequencies, sensitivity analyses used the GWAS-side allele frequencies. Full methods, results, and interpretation are provided in Supplementary Note 1 and Supplementary Table S7.

## 3. Results

### 3.1 Instrument characteristics

Across six CHD subtype exposures, genome-wide significant instrument counts ranged from 0 (ToF) to 98 (ASD) (Table 1). After streaming extraction of 155 instruments and matching to the outcome GWAS (125–137 matched per outcome), harmonisation with palindromic-SNP removal and F > 10 filtering left 10–78 instruments per pair. All retained instruments had F > 10 (minimum F 29.9–33.1 in the aortopathy anchors).

### 3.2 Cross-cohort primary analysis

Table 2 reports IVW OR with 95% CI and P-value for all 12 exposure–outcome pairs, alongside MR-Egger intercept, weighted median, Cochran’s Q, Bonferroni P, and Steiger direction; the complete results for every exposure–outcome pair across all MR methods, including the abdominal-aneurysm analysis, are provided in Supplementary Table S1. The primary comparison is whether BAV and LVOTO/CoA show a specific, robust effect on AF that exceeds their effect on HF, in independent cohorts.

**Table 2.** Cross-cohort primary MR estimates (FinnGen exposures × independent GWAS Catalog harmonised outcomes). IVW OR per genetically determined CHD subtype, 95% CI. n = SNPs retained after harmonisation and F filtering. Steiger directionality was recomputed using FinnGen R13 endpoint-specific case counts with liability-scale SNP– trait correlations from get_r_from_lor() (Methods). †The overall-CHD → thoracic-aneurysm estimate is a broad-phenotype result with near-perfect homogeneity (Q p=1.000) and is not interpreted as a subtype-level causal effect (§3.4).

| Exposure → Outcome | n SNP | IVW OR (95% CI) | P | Egger intercept P | Q P | WMed OR | Bonferroni P | Steiger direction |
|---|---|---|---|---|---|---|---|---|
| BAV → atrial fibrillation | 14 | 1.046 (1.031–1.062) | 1.7×10<sup>−9</sup> | 0.031 | 0.852 | 1.038 | 2.1×10<sup>−8</sup> | TRUE |
| LVOTO → atrial fibrillation | 19 | 1.065 (1.042–1.088) | 8.5×10<sup>−9</sup> | 0.353 | 0.730 | 1.053 | 1.0×10<sup>−7</sup> | TRUE |
| ASD → atrial fibrillation | 78 | 1.014 (1.001–1.028) | 0.041 | 0.006 | 1.1×10<sup>−31</sup> | 0.996 | 0.488 | TRUE |
| Overall CHD → atrial fibrillation | 62 | 1.295 (1.257–1.334) | 5.9×10<sup>−66</sup> | 4.4×10<sup>−7</sup> | 4.46×10<sup>−10</sup> | 1.309 | 7.1×10<sup>−65</sup> | TRUE |
| BAV → heart failure | 10 | 1.024 (1.004–1.045) | 0.022 | 0.228 | 0.991 | 1.024 | 0.259 | TRUE |
| LVOTO → heart failure | 14 | 1.034 (1.005–1.065) | 0.022 | 0.558 | 0.998 | 1.029 | 0.266 | TRUE |
| ASD → heart failure | 55 | 1.000 (0.989–1.012) | 0.974 | 1.9×10<sup>−5</sup> | 0.003 | 0.988 | 1 | TRUE |
| Overall CHD → heart failure | 50 | 1.198 (1.170–1.227) | 1.7×10<sup>−50</sup> | 0.219 | 0.974 | 1.209 | 2.1×10<sup>−49</sup> | TRUE |
| BAV → thoracic aortic aneurysm | 10 | 0.935 (0.841–1.041) | 0.221 | 0.512 | 1.000 | 0.924 | 1 | TRUE |
| LVOTO → thoracic aortic aneurysm | 14 | 0.889 (0.763–1.036) | 0.132 | 0.431 | 1.000 | 0.907 | 1 | TRUE |
| ASD → thoracic aortic aneurysm | 61 | 1.022 (0.976–1.072) | 0.353 | 0.397 | 1.000 | 0.978 | 1 | TRUE |
| Overall CHD → thoracic aortic aneurysm† | 50 | 1.559 (1.372–1.770) | 8.0×10<sup>−12</sup> | 0.314 | 1.000 | 1.737 | 9.7×10<sup>−11</sup> | TRUE |

### 3.3 Subtype-resolved AF effects and outcome specificity

The two aortopathy-prone subtypes showed small but statistically robust effects on AF. BAV conferred an OR of 1.046 (1.031–1.062; IVW p=1.7×10⁻⁹; Bonferroni p=2.1×10⁻⁸) and LVOTO/CoA an OR of 1.065 (1.042–1.088; IVW p=8.5×10⁻⁹; Bonferroni p=1.0×10⁻⁷). Both estimates were concordant across weighted median (1.038 and 1.053) and weighted mode (1.029 and 1.0442), showed no heterogeneity (Q p=0.852 and 0.730), and had correct Steiger direction and stable leave-one-out ranges. For BAV→AF the MR-Egger intercept was borderline (p=0.031), whereas for LVOTO→AF it was not significant (p=0.353); the weighted-median estimates remained close to IVW, so the effects are not driven by a single pleiotropic outlier, and MR-PRESSO detected no outlier SNP in either pair.

Outcome specificity was preserved: HF effects for the same subtypes were smaller and did not survive Bonferroni correction (BAV OR 1.024, p=0.022, Bonferroni p=0.259; LVOTO OR 1.034, p=0.022, Bonferroni p=0.266). The hierarchy AF > HF for BAV and LVOTO is consistent with a subtype-specific arrhythmic predisposition rather than a generic effect on all late complications. Broad categories behaved differently: overall CHD showed large effects on AF (OR 1.295) and HF (OR 1.198), while ASD→AF was borderline and non-robust (OR 1.014, nominal p=0.041, Bonferroni p=0.488, with pronounced heterogeneity Q p=1.1×10⁻³¹ and a significant Egger intercept p=0.006). Thus the subtype-resolved AF signal is specific to BAV and LVOTO/CoA and is not reproduced by the ASD subtype.

### 3.4 Non-replication of the aortopathy signal and interpretation of the broad-phenotype estimate

The aortopathy-anchor estimates from the same-cohort pilot (BAV → aneurysm OR 1.45; LVOTO OR 1.77) were not reproduced with independent outcomes: BAV → thoracic aortic aneurysm gave OR 0.935 (0.841–1.041; p=0.221) and LVOTO OR 0.889 (0.763–1.036; p=0.132), both with confidence intervals including the null and excluding the same-cohort point estimates. Two features could in principle temper this negative finding: the thoracic aneurysm GWAS has only 1,351 cases, and its phenotype (thoracic) is not identical to the FinnGen I9_AORTANEUR endpoint (ICD-10 I71, predominantly abdominal). We addressed both directly. First, power calculations showed that limited statistical power is not the explanation: the analysis had 80% power to detect ORs ≥1.17 (BAV) and ≥1.24 (LVOTO), >99.9% power to detect the same-cohort estimates (OR 1.45 and 1.77), and the cross-cohort estimates differed from the same-cohort ones by >8 standard errors (p<10⁻¹⁵). Second, to test the phenotype-mismatch explanation we repeated the analysis using an independent, harmonised abdominal aortic aneurysm GWAS (GCST90475994; 16,289 cases / 427,111 controls). BAV and LVOTO were again null (BAV OR 0.987, 0.960–1.015, p=0.37; LVOTO OR 0.981, 0.942–1.021, p=0.35), with no heterogeneity (Cochran’s Q p≈1), no directional pleiotropy (MR-Egger intercept p>0.2), and correct Steiger direction. The same-cohort aortopathy signal is therefore most consistent with sample overlap and winner’s curse, and non-replication here constitutes robust evidence against a large subtype-specific aortopathy effect (while not excluding small effects, OR≲1.1).

The largest cross-cohort point estimate—overall CHD → thoracic aortic aneurysm, OR 1.559 (1.372–1.770; p=8.0×10⁻¹²)—is not interpreted as a subtype-level causal effect and is flagged for three reasons. First, it derives from a broad “circulatory congenital malformation” phenotype rather than a defined subtype, and its magnitude exceeds every subtype-specific estimate, a pattern more consistent with shared developmental liability or residual heterogeneity than with a single causal pathway. Second, it showed near-perfect homogeneity (Q p=1.000), an atypical pattern that, alongside a non-significant Egger intercept (p=0.314), does not discriminate pleiotropy well and warrants caution: with 50 instruments, Q p=1.000 implies less between-instrument dispersion than sampling error predicts, consistent with highly correlated instruments or over-precise effect estimates, which further argues against interpreting this as a subtype-level causal effect. Third, its direction is opposite to the subtype-level results (BAV and LVOTO were both null for aneurysm). We note that an initial Steiger test using a quantitative-trait approximation had flagged this pair as directionally inconsistent; recomputation with endpoint-specific case counts and liability-scale SNP–trait correlations (get_r_from_lor()) reversed that flag (Steiger p=8.8×10⁻³¹, correct direction TRUE). We therefore do not rest the exclusion of this pair on directionality, but on its broad phenotype, anomalous homogeneity, and inconsistency with the subtype analyses. All Steiger tests in the final analysis used FinnGen R13 endpoint-specific case counts (BAV 1,356; LVOTO 4,050; ASD 1,443; overall CHD 6,216; total N=500,186) and get_r_from_lor()-derived correlations; the primary AF conclusions were unchanged by this correction.

### 3.5 Same-cohort analysis as a bias illustration

To quantify overlap bias, we repeated the analyses with FinnGen outcomes (Table 3). The same-cohort estimates were systematically larger: BAV → aneurysm OR 1.45 versus 0.935 cross-cohort; LVOTO → aneurysm OR 1.77 versus 0.889; BAV → AF OR 1.12 versus 1.046; LVOTO → AF OR 1.17 versus 1.065. Broad categories were most inflated (overall CHD → aneurysm OR 1.95; → AF OR 1.85), consistent with extensive shared controls and winner’s curse. Per the prespecified adjudication rule (§2.5), these values are used only as a gauge of bias magnitude and are not causal estimates.

**Table 3.** Within-FinnGen (same-cohort) IVW estimates, presented solely as a bias illustration (from E_pilot_results.txt).

| Exposure → Outcome | n | IVW OR (95% CI) | Cross-cohort OR | Egger intercept P | Q P |
|---|---|---|---|---|---|
| BAV → aortic aneurysm | 15 | 1.45 (1.41–1.50) | 0.935 | 0.058 | 0.99 |
| LVOTO → aortic aneurysm | 21 | 1.77 (1.70–1.85) | 0.889 | <1×10<sup>−300</sup> | 6×10<sup>−4</sup> |
| BAV → AF | 15 | 1.12 (1.10–1.14) | 1.046 | 6×10<sup>−7</sup> | 0.21 |
| LVOTO → AF | 21 | 1.17 (1.14–1.20) | 1.065 | 0.014 | 0.02 |
| BAV → HF | 15 | 1.05 (1.03–1.07) | 1.024 | 2×10<sup>−11</sup> | 1.00 |
| LVOTO → HF | 21 | 1.08 (1.05–1.11) | 1.034 | 0.029 | 1.00 |
| Overall CHD → aneurysm | 69 | 1.95 (1.88–2.01) | 1.559 | <1×10<sup>−300</sup> | 1×10<sup>−12</sup> |
| Overall CHD → AF | 69 | 1.85 (1.82–1.89) | 1.295 | 6×10<sup>−7</sup> | 1×10<sup>−12</sup> |
| ASD → AF | 95 | 1.06 (1.05–1.07) | 1.014 | 0.89 | 1.00 |
| ASD → HF | 95 | 0.98 (0.97–0.99) | 1.000 | 0.98 | 2×10<sup>−12</sup> |

### 3.6 Sensitivity analyses

For MR-PRESSO (MRPRESSO v1.0), the global heterogeneity test was estimable and non-significant for the primary subtype pairs—BAV→AF P=0.86 and LVOTO→AF P=0.80 (14 and 19 instruments, respectively, sufficient for the test)—and the outlier test detected no outlier SNP in either pair, indicating that the AF estimates were not driven by single influential variants. Here “non-significant” denotes an estimable global test yielding P>0.05, not a test that could not be performed for lack of instruments; for the abdominal-aneurysm sensitivity analysis the global test was not evaluated because that GWAS lacked the standard-error field needed for outlier detection (Supplementary Table S2). For completeness, the global test was significant (P<0.01) for ASD→AF and overall-CHD→AF, although the outlier test still identified no single outlier; this pattern is consistent with diffuse heterogeneity across many instruments rather than a discrete pleiotropic variant, and reinforces our decision not to interpret these broad or non-focal exposures as robust findings. Leave-one-out estimates for BAV→AF and LVOTO→AF were stable. The main interpretive cautions are (i) the borderline Egger intercept for BAV→AF, (ii) pronounced heterogeneity for ASD→AF (Q p=1.1×10⁻³¹), and (iii) the broad-phenotype overall CHD→TAA estimate with anomalous homogeneity (§3.4). These are handled in the primary interpretation rather than hidden.

## 4. Discussion

### 4.1 Principal findings

In independent, non-overlapping cohorts, genetically determined BAV and LVOTO/CoA conferred a small but robust and subtype-specific increase in late AF risk (OR 1.046 and 1.065; Bonferroni p=2.1×10⁻⁸ and 1.0×10⁻⁷), with smaller HF effects that did not survive multiple-testing correction. In parallel, the large aortopathy effects seen in same-cohort analyses were not replicated: point estimates were below unity and their intervals excluded the same-cohort values. The AF odds ratios reported here are relative measures and should not be read as absolute risk increases. The study therefore makes two contributions: a clinically usable, subtype-resolved AF result, and an empirical quantification of how badly same-cohort design can mislead MR of rare congenital exposures. Because summary data cannot separate sample overlap from winner’s curse, the 1.45→0.935 and 1.77→0.889 gaps should be read as upper-bound estimates of their joint bias rather than as a pure overlap-bias magnitude.

### 4.2 Interpretation in context

The subtype-level AF effect we observed is compatible in direction with the only prior broad-category MR, which reported congenital great-vessel anomalies increasing AF risk (OR ≈ 1.07)^[8]^; our BAV and LVOTO estimates are of comparable magnitude and provide subtype-level corroboration rather than a contradictory signal. What this study adds is subtype resolution plus an independent-cohort design: the broad category averages across heterogeneous subtypes, whereas the aortopathy-prone subtypes carry a distinct, reproducible arrhythmic liability. We also note the conceptual reverse direction—genetically determined aortic diameter increasing LVOT (including CoA) CHD risk—established by Tcheandjieu et al.^[19]^ in Circulation: Genomic and Precision Medicine; our forward-direction, subtype-resolved AF result complements that work and shifts the clinical question from aortic surveillance alone to combined aortic and arrhythmic surveillance.

Instrument architecture also helps explain the contrast between subtypes. The ASD instruments (n=78) are dispersed across many loci and show pronounced heterogeneity (Q p=1.1×10⁻³¹) and a significant Egger intercept (p=0.006), whereas the BAV and LVOTO instruments, although concentrated at a single 1p22 locus, yield low heterogeneity and stable, concordant estimates. Signal reliability in MR therefore depends on instrument architecture as well as effect magnitude: a dispersed multi-locus instrument set is more exposed to heterogeneous and pleiotropic effects, while a focused developmental locus can give a cleaner—albeit biologically narrower— estimate. This contrast reinforces the value of subtype resolution rather than undermining it.

A further observation concerns the magnitude of same-cohort inflation across outcomes. The aortopathy estimates were inflated roughly 1.55-fold (1.45/0.935) and 1.99-fold (1.77/0.889), whereas the AF estimates were inflated only about 1.07-fold (1.12/1.046) and 1.10-fold (1.17/1.065). This gradient is compatible with the precision difference between the independent outcome GWAS: the thoracic-aneurysm study has only 1,351 cases, so shared controls and winner’s curse can more readily distort a noisy point estimate, while the AF study (60,620 cases) yields a more stable outcome association that dilutes the relative impact of overlap. We present this as an observational hypothesis—that independent-outcome sample size modulates the apparent magnitude of same-cohort bias—which should be tested in future methodological work rather than treated as an established mechanism here.

### 4.3 Biological interpretation of the BAV/LVOTO instruments

A reviewer-relevant step for this journal is to make the instruments biologically interpretable rather than merely statistically significant. We annotated all BAV and LVOTO lead instruments against the FinnGen R13 summary statistics (nearest-gene field; Supplementary Table S5). This revealed that the instruments are highly concentrated in a single 1p22 locus annotated to LINC01708, adjacent to PALMD (34 of the 36 rsID-bearing instruments: BAV 15/15, LVOTO 19/21); this is an established BAV and aortic-root-size association signal. We next asked whether this GWAS signal is driven by PALMD expression by performing Bayesian colocalization (coloc.abf) between each exposure and PALMD cis-eQTLs from three GTEx tissues in the eQTL Catalogue (aorta, atrial appendage, left ventricle; ∼3,850 shared SNPs per pair; Supplementary Table S7, Supplementary Note 1). All six pairs gave PP4 < 0.003 for a shared causal variant, with the posterior mass instead on H3 (two distinct causal variants; PP3 ≈ 0.99– 1.00 in atrium and left ventricle) or H1 (a GWAS signal with no corresponding eQTL signal; PP1 ≈ 0.97 in aorta). The BAV/LVOTO lead SNPs (e.g., rs11166276, rs6702619) were not associated with PALMD expression in any tissue (all P > 0.05), and the GWAS lead peak (∼99.56 Mb) lies ∼100 kb from the strongest PALMD eQTL peak (∼99.66 Mb), indicating two separable signals rather than one shared causal variant. These data therefore do not support PALMD as the causal effector gene at this locus in the tissues available; the effector may instead be LINC01708 itself or a regulatory element not captured by GTEx eQTLs. We stress that this does not overturn the MR finding— colocalization addresses effector-gene identity, not the validity of the instrument– outcome association—and that GTEx lacks an aortic-valve-specific tissue, so a valve-restricted eQTL effect cannot be excluded. Consistent with this, nearly all instruments derive from one locus, so the AF signal is biologically anchored but is not corroborated by multiple independent developmental pathways (e.g., NOTCH1, GATA/NKX2-5, TGF-β); nearest-gene assignment does not establish the causal gene, and an F-statistic alone does not establish clinical interpretability. The complete negative colocalization analysis, including prespecified decision rules and limitations, is reported as Supplementary Note 1 to make this inference fully auditable. With respect to the borderline MR-Egger intercept for BAV→AF (p=0.031), we note that concordance of the weighted-median estimate argues against a single pleiotropic outlier but cannot exclude correlated pleiotropy—multiple instruments acting through a shared unmeasured pathway—which neither the outlier test nor colocalization at one locus can fully rule out. We therefore interpret the BAV→AF effect as robust to outlier-driven pleiotropy but not established as a single-pathway mechanism. A pattern-level observation is informative here: the AF estimates are Bonferroni-robust (p=2.1×10⁻⁸ and 1.0×10⁻⁷ for BAV and LVOTO), whereas the HF estimates for the same exposures are not (Bonferroni p=0.259 and 0.266), and the LVOTO→AF Egger intercept is non-significant (p=0.353). This outcome specificity is more compatible with a subtype-linked arrhythmic liability than with a single shared pleiotropic pathway driving all cardiovascular endpoints, although it is suggestive rather than definitive and the exclusion restriction remains untestable with summary data. We did not apply MR-Egger SIMEX, which is not implemented in TwoSampleMR, and state this boundary explicitly.

### 4.4 Strengths

Key strengths include (i) a cross-cohort, non-overlapping design appropriate for rare congenital exposures, with harmonised outcome GWAS; (ii) subtype resolution rather than a broad CHD label, isolating BAV and LVOTO/CoA effects on AF; (iii) explicit quantification of same-cohort bias using a prespecified adjudication rule, turning a common methodological liability into a reported result; (iv) a multi-method MR framework (IVW, Egger, weighted median/mode, MR-PRESSO, Steiger, leave-one-out) with Bonferroni correction; and (v) transparent handling of rare-disease power limits (ToF excluded with a stated structural reason; VSD flagged as weak-evidence). We also note that the negative effector-gene colocalization was prespecified, fully reported, and separated from the primary causal inference (Supplementary Note 1), which we view as a methodological strength rather than a limitation.

### 4.5 Limitations

First, CHD is etiologically heterogeneous; even subtype-defined instruments capture only part of the genetic liability, and developmental pleiotropy may violate the exclusion restriction (the borderline BAV→AF Egger intercept is a case in point). In particular, correlated pleiotropy cannot be excluded by our robustness analyses, so the AF estimates should not be read as free of all directional pleiotropy. Second, the late-complication framing means CHD exposures are paediatric while AF/HF manifest in adulthood; without an age lower bound, estimates reflect lifetime cumulative liability, not time-window-specific risk. Third, rare-subtype instruments are few (BAV n=14, LVOTO n=19 after harmonisation for AF), so effects are modest and confidence intervals, though excluding the null, are not wide enough to rule out smaller true effects in other ancestries. Fourth, the thoracic aneurysm GWAS has only 1,351 cases, but we complemented it with power calculations and an independent abdominal aortic aneurysm GWAS (GCST90475994, 16,289 cases), which together argue against power or phenotype mismatch as explanations for non-replication (§3.4); we cannot, however, exclude small aortopathy effects (OR≲1.1). Fifth, Steiger directionality was computed with endpoint-specific case counts and liability-scale correlations (get_r_from_lor()); an initial quantitative-trait approximation had misclassified one broad-phenotype pair, which we corrected, and the AF conclusions were unaffected. Sixth, harmonised outcome EAF values were frequently missing, constraining palindromic-SNP resolution. Seventh, FinnGen is predominantly European; generalizability is untested. Eighth, the BAV/LVOTO instruments are concentrated in a single 1p22 LINC01708/PALMD locus (§4.3, Table S5), so the subtype AF signal is anchored to one developmental locus rather than replicated across multiple independent loci; this limits the biological breadth of the inference and means the estimates should not be read as the net effect of all BAV/LVOTO-related pathways. Colocalization further showed that this GWAS signal does not share a causal variant with PALMD eQTLs in the GTEx tissues available (all PP4 < 0.003; Table S7, Supplementary Note 1), so the effector gene at this locus remains unresolved and the valve-development mechanism is not established by these data. Finally, MR estimates a lifetime population-average effect of genetic liability and should not be read as the effect of surgical correction or current management.

### 4.6 Conclusion

In independent, non-overlapping cohorts, genetically determined BAV and LVOTO/CoA confer a modest but robust increase in AF risk (OR 1.046 and 1.065; Bonferroni p=2.1×10⁻⁸ and 1.0×10⁻⁷), with smaller heart-failure effects that do not survive multiple-testing correction, whereas the large aortopathy effects seen in same-cohort analyses were not replicated. Same-cohort MR can therefore materially overstate subtype-specific effects, and overlap bias should be quantified rather than assumed away in two-sample MR of rare congenital exposures. Subtype-aware arrhythmia surveillance in BAV and CoA survivors, and cross-cohort replication as a reporting standard, are the practical implications.

## Data Availability

All summary statistics used in this study are publicly available. Exposure instruments were obtained from FinnGen R13 (https://www.finngen.fi; endpoints Q17_CONGEN_MALFO_CIRCULATO_SYSTEM, Q17_ASD, Q17_VENTRICULAR_SEPTAL_DEFECT, Q17_LVOTO_BROAD, Q17_BICUSP_AORTIC_VALVE, and bias-analysis endpoints I9_AORTANEUR, I9_AF, I9_HEARTFAIL, I9_VTE). Outcomes were GWAS Catalog harmonised summary statistics (https://www.ebi.ac.uk/gwas): GCST90027266 (thoracic aortic aneurysm), GCST90475994 (abdominal aortic aneurysm), GCST006414 (atrial fibrillation), and GCST009541 (heart failure). eQTL data were from the eQTL Catalogue GTEx v8 datasets QTD000131 (aorta), QTD000251 (atrial appendage), and QTD000256 (left ventricle). Analysis code is provided as supplementary material.

https://www.ebi.ac.uk/eqtl?eQTL_Catalogue?

https://www.finngen.fi

https://www.ebi.ac.uk/gwas

## Nonstandard Abbreviations and Acronyms

CHD: congenital heart disease
MR: Mendelian randomization
IVW: inverse-variance weighted
BAV: bicuspid aortic valve
LVOTO: left-ventricular outflow tract obstruction
CoA: coarctation of the aorta
ASD: atrial septal defect
VSD: ventricular septal defect
ToF: tetralogy of Fallot
AF: atrial fibrillation
HF: heart failure
TAA: thoracic aortic aneurysm
VTE: venous thromboembolism
GWAS: genome-wide association study
SNP: single-nucleotide polymorphism
TOP: Transparency and Openness Promotion Clinical Perspective

## Declarations

Data availability. All summary statistics are publicly available. Exposure instruments: FinnGen R13 (https://www.finngen.fi; endpoints Q17_CONGEN_MALFO_CIRCULATO_SYSTEM, Q17_ASD, Q17_VENTRICULAR_SEPTAL_DEFECT, Q17_LVOTO_BROAD, Q17_BICUSP_AORTIC_VALVE, and bias-analysis endpoints I9_AORTANEUR, I9_AF, I9_HEARTFAIL, I9_VTE). Outcomes: GWAS Catalog (https://www.ebi.ac.uk/gwas) harmonised summary statistics GCST90027266 (thoracic aortic aneurysm), GCST90475994 (abdominal aortic aneurysm), GCST006414 (atrial fibrillation), and GCST009541 (heart failure). eQTL data: eQTL Catalogue GTEx v8 datasets QTD000131 (aorta), QTD000251 (atrial appendage), and QTD000256 (left ventricle). Analysis code is provided as supplementary material (E_prepare_cross_cohort.py, E_extract_outcome.awk, E_extract_AAA.py, E_R_cross_cohort.R, E_R_one_outcome.R, E_R_steiger_fix.R, E_coloc_extract_gwas.py, E_coloc_extract_eqtl.py, E_coloc_align.R, E_coloc_run.R).

## AI-assisted drafting disclosure

This manuscript was prepared with the assistance of an AI writing tool. All factual claims, statistical results, and citations were verified by the authors against primary sources, and AI use is disclosed in accordance with the target journal’s policy.

## Supplementary Material

- Supplementary Note 1. Bayesian colocalization of the 1p22 BAV/LVOTO signal with PALMD cis-eQTLs: a negative result and its methodological implications.

- Table S1. Full MR results (all exposure–outcome pairs, all methods): E_results_cross_cohort.csv, E_results_cross_cohort_methods.csv; abdominal-aneurysm analysis: E_results_AAA.csv, E_Table_AAA.md.

- Table S2. MR-PRESSO outputs (no outliers detected in any pair).

- Table S3. Leave-one-out analyses.

- Table S4. Steiger directionality outputs (E_results_steiger_fixed.csv; recomputed with get_r_from_lor() and endpoint-specific case counts).

- Table S5. Instrument-to-gene biological annotation for BAV and LVOTO lead instruments (E_S5_instrument_annotation.tsv/.md; nearest gene from FinnGen R13; 34/36 rsID-bearing instruments at the 1p22 LINC01708/PALMD locus; table note documents the full instrument flow from 16/22 in FinnGen to 14/19 in the final harmonised AF analysis).

- Table S6. Methods note — reconstruction of standard errors for the abdominal aortic aneurysm GWAS. GCST90475994 reported no standard_error field; for each instrument SE[log(OR)] was recovered from the 95% CI as SE = [ln(CI_upper) − ln(CI_lower)]/3.92, with beta = ln(OR). These values were used only for the abdominal-aneurysm sensitivity analysis and did not affect the primary AF results.

- Table S7. 1p22–PALMD colocalization (E_S7_colocalisation.csv/.md; coloc.abf v5.2.3; BAV/LVOTO × aorta/atrium/left-ventricle cis-eQTLs; ∼3,850 SNPs per pair; all PP4 < 0.003 — no support for a shared causal variant).

---

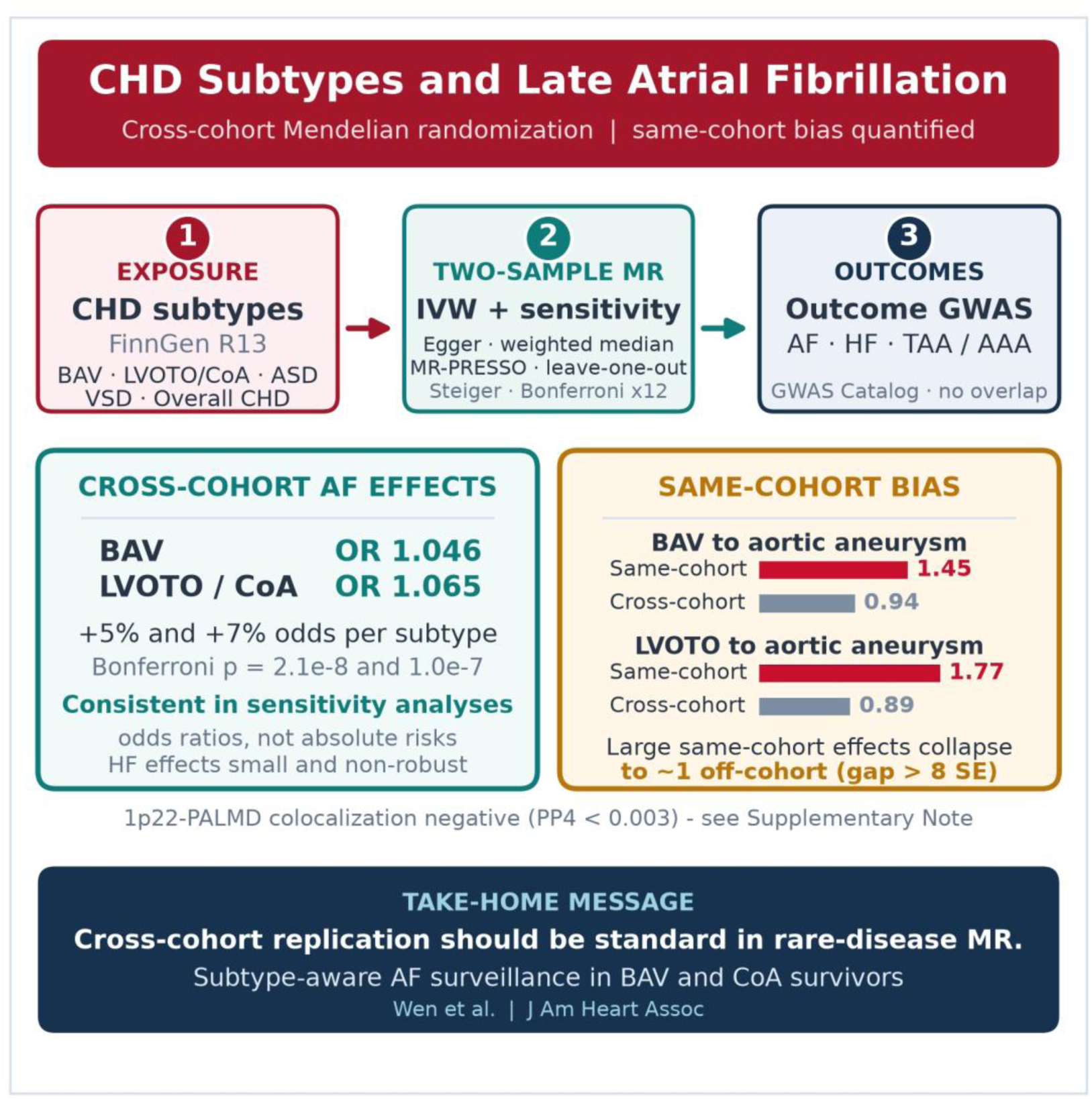

## Supplementary Note 1

### Bayesian colocalization of the 1p22 BAV/LVOTO signal with PALMD cis-eQTLs: a prespecified negative result and its methodological implications

#### 1. Why this analysis was performed

Supplementary Table S5. showed that 34 of 36 genome-wide significant BAV/LVOTO lead instruments map to a single 1p22 locus annotated to LINC01708, adjacent to PALMD. Because a nearest-gene annotation is only indicative, we asked whether the 1p22 GWAS signal and PALMD expression are driven by the same causal variant. This is a question about effector-gene identity at the instrument locus, not a test of the primary CHD-subtype → atrial fibrillation (AF) causal estimate. We prespecified the decision rule before examining the output: PP4 ≥ 0.8 would support a shared causal variant (and hence PALMD as the likely effector); PP4 < 0.5 would not support colocalization. We report the negative result in full because it defines the biological boundary of the MR inference and makes the interpretation auditable.

#### 2. Data and methods

- GWAS side. FinnGen R13 summary statistics for BAV (Q17_BICUSP_AORTIC_VALVE) and LVOTO (Q17_LVOTO_BROAD), restricted to chr1:99.0–100.2 Mb (GRCh38) around the lead-signal peak, extracted by tabix-style indexed range query (E_coloc_extract_gwas.py, E_tabix_query.py).

- eQTL side. PALMD (ENSG00000099260) cis-eQTLs from the eQTL Catalogue GTEx v8 datasets: aorta QTD000131, atrial appendage QTD000251, and left ventricle QTD000256 (E_coloc_extract_eqtl.py).

- Harmonisation. Variants were matched by rsid; effect alleles were aligned so that beta signs referred to the same allele; palindromic and ambiguous variants were removed. This left 3,853–3,869 shared SNPs per pair (E_coloc_align.R).

- Colocalization. coloc.abf (v5.2.3) with default priors p1 = p2 = 1×10⁻⁴ and p12 = 1×10⁻⁵; sample sizes N = 500,186 (GWAS) and N = 500,000 (eQTL). Posterior probabilities for H0–H4 were extracted from res$summary (E_coloc_run.R). Because the harmonised outcome files lacked effect-allele frequencies, the analysis used GWAS-side allele frequencies as the MAF input under coloc.abf’s single-causal-variant-per-trait assumption, without an external LD matrix. We therefore interpret PP4 conservatively.

#### 3. Results

All six exposure–tissue pairs failed the prespecified colocalization criterion: PP4 ranged from 2.6×10⁻¹⁰ to 2.8×10⁻³, far below the 0.8 threshold, and PP4/PP3 was ≤ 0.121 in every pair.

| Pair | nSNP | PP.H0 | PP.H1 | PP.H2 | PP.H3 | PP.H4 (PP4) | PP4/PP3 | Verdict |
|---|---|---|---|---|---|---|---|---|
| BAV × aorta | 3,869 | 4.054×10<sup>-6</sup> | 0.9742 | 9.567×10<sup>-8</sup> | 0.02299 | 0.002786 | 0.121 | No support |
| BAV × atrium | 3,859 | 3.879×10<sup>-8</sup> | 0.009323 | 4.122×10<sup>-6</sup> | 0.9906 | 4.051×10<sup>-5</sup> | 4.09×10<sup>-5</sup> | No support |
| BAV × left ventricle | 3,853 | 2.695×10<sup>-20</sup> | 6.477×10<sup>-15</sup> | 4.161×10<sup>-6</sup> | 1.000 | 2.615×10<sup>-8</sup> | 2.62×10<sup>-8</sup> | No support |
| LVOTO × aorta | 3,869 | 6.474×10<sup>-8</sup> | 0.9743 | 1.528×10<sup>-9</sup> | 0.02299 | 0.002749 | 0.120 | No support |
| LVOTO × atrium | 3,859 | 6.196×10<sup>-10</sup> | 0.009324 | 6.583×10<sup>-8</sup> | 0.9906 | 3.998×10<sup>-5</sup> | 4.04×10<sup>-5</sup> | No support |
| LVOTO × left ventricle | 3,853 | 4.304×10<sup>-22</sup> | 6.477×10<sup>-15</sup> | 6.646×10<sup>-8</sup> | 1.000 | 2.804×10<sup>-10</sup> | 2.80×10<sup>-10</sup> | No support |

Two features reinforce the posterior result. First, the lead BAV/LVOTO SNPs (e.g., rs11166276 and rs6702619) were not associated with PALMD expression in any of the three tissues (all P > 0.05). Second, the GWAS lead peak (∼99.56 Mb) and the strongest PALMD eQTL peak (∼99.66 Mb) are separated by approximately 100 kb, consistent with two distinct regulatory signals rather than one shared causal variant. The posterior mass lay on H3 (two distinct causal variants; PP3 ≈ 0.99–1.00 in atrium and left ventricle) or on H1 (a GWAS signal without a matched eQTL signal; PP1 ≈ 0.97 in aorta).

#### 4. Interpretation

These data do not support PALMD as the causal effector gene at the 1p22 locus in the tissues available. The effector may be LINC01708 itself or a regulatory element not captured by GTEx eQTLs. Importantly, this negative result does not overturn the MR finding: colocalization addresses which gene transmits a locus-level signal, whereas MR tests the exposure–outcome association through the instrument set. A locus can be a valid instrument for BAV/LVOTO liability even when its effector gene is unresolved. Consistent with this, nearly all instruments derive from one locus, so the AF signal is biologically anchored but is not corroborated by multiple independent developmental pathways (e.g., NOTCH1, GATA/NKX2-5, TGF-β).

#### 5. Methodological transparency

We highlight four reasons for reporting this analysis as a standalone note. (i) The decision rule was fixed before the result was inspected, so the negative finding is not a post hoc reinterpretation. (ii) The analysis replaces a suggestive nearest-gene annotation with an explicit statistical test, preventing an unsupported mechanism from entering the Discussion as fact. (iii) The primary causal inference and the effector-gene inference are reported separately, so readers can judge them on their own evidence. (iv) Full posterior probabilities, harmonisation code, and colocalization scripts are supplied (E_coloc_extract_gwas.py, E_coloc_extract_eqtl.py, E_coloc_align.R, E_coloc_run.R, E_coloc_sensitivity.R; full results in Supplementary Table S7), permitting independent reproduction. We regard this as a strength of the study rather than a qualification to be hidden.

#### 6. Limitations of this note

GTEx does not include an aortic-valve-specific tissue; the aorta, atrial appendage, and left ventricle are surrogate tissues, and a valve-restricted eQTL effect cannot be excluded. eQTL power is limited in aorta (n ≈ 224), so a low PP4 is not proof that PALMD has no regulatory role at this locus. coloc.abf assumes at most one causal variant per trait in the region and was run without an external LD matrix using GWAS-side allele frequencies; in the presence of multiple causal signals, PP4 can be attenuated. Finally, colocalization does not assess whether the instruments satisfy the MR exclusion restriction; that question is addressed by the MR sensitivity analyses in the main text.

##### 6.1 Prior sensitivity analysis

Because coloc.abf was run without an external LD matrix, we tested whether the negative conclusion is sensitive to the choice of the p12 prior (the prior probability that a variant is causal for both traits). We repeated all six pairs with p1 = p2 = 10⁻⁴ fixed and p12 set to 10⁻⁶ and 10⁻⁴, alongside the default p12 = 10⁻⁵. The posterior ranking was unchanged under every prior: H3 (two distinct causal variants) remained dominant in atrium and left ventricle (PP3 ≈ 0.99–1.00), and H1 (GWAS signal without a matched eQTL) dominated in aorta. PP4 for a shared causal variant never approached the 0.8 support threshold; even under the most H4-favourable prior (p12 = 10⁻⁴), the maximum PP4 was 0.027, reached independently in BAV × aorta (0.0272) and LVOTO × aorta (0.0268), and PP4 in the other four pairs remained below 5×10⁻⁴. The conclusion that the 1p22 GWAS signal and PALMD cis-eQTLs do not share a causal variant is therefore robust to the p12 prior.

| Pair | Prior | nSNP | PP.H3 | PP.H4 |
|---|---|---|---|---|
| BAV × aorta | default (p12=10<sup>-5</sup>) | 3,869 | 0.0230 | 0.00279 |
| BAV × aorta | p12=10<sup>-6</sup> | 3,869 | 0.0231 | 0.00028 |
| BAV × aorta | p12=10<sup>-4</sup> | 3,869 | 0.0224 | 0.0272 |
| BAV × atrium | default (p12=10<sup>-5</sup>) | 3,859 | 0.9906 | 4.05×10<sup>-5</sup> |
| BAV × atrium | p12=10<sup>-6</sup> | 3,859 | 0.9907 | 4.05×10<sup>-6</sup> |
| BAV × atrium | p12=10<sup>-4</sup> | 3,859 | 0.9903 | 4.05×10<sup>-4</sup> |
| BAV × left ventricle | default (p12=10<sup>-5</sup>) | 3,853 | 1.000 | 2.62×10<sup>-8</sup> |
| BAV × left ventricle | p12=10<sup>-6</sup> | 3,853 | 1.000 | 2.62×10<sup>-9</sup> |
| BAV × left ventricle | p12=10<sup>-4</sup> | 3,853 | 1.000 | 2.62×10<sup>-7</sup> |
| LVOTO × aorta | default (p12=10<sup>-5</sup>) | 3,869 | 0.0230 | 0.00275 |
| LVOTO × aorta | p12=10<sup>-6</sup> | 3,869 | 0.0231 | 0.00028 |
| LVOTO × aorta | p12=10<sup>-4</sup> | 3,869 | 0.0224 | 0.0268 |
| LVOTO × atrium | default (p12=10<sup>-5</sup>) | 3,859 | 0.9906 | 4.00×10<sup>-5</sup> |
| LVOTO × atrium | p12=10<sup>-6</sup> | 3,859 | 0.9907 | 4.00×10<sup>-6</sup> |
| LVOTO × atrium | p12=10<sup>-4</sup> | 3,859 | 0.9903 | 4.00×10<sup>-4</sup> |
| LVOTO × left ventricle | default (p12=10<sup>-5</sup>) | 3,853 | 1.000 | 2.80×10<sup>-10</sup> |
| LVOTO × left ventricle | p12=10<sup>-6</sup> | 3,853 | 1.000 | 2.80×10<sup>-11</sup> |
| LVOTO × left ventricle | p12=10<sup>-4</sup> | 3,853 | 1.000 | 2.80×10<sup>-9</sup> |

#### 7. Reproducibility

All inputs are public: FinnGen R13 (https://www.finngen.fi) and the eQTL Catalogue (https://www.ebi.ac.uk/eqtl/; GTEx v8 QTD000131, QTD000251, QTD000256). Analysis scripts and posterior probabilities are provided as supplementary material; numeric results are reproduced in Supplementary Table S7 (E_S7_colocalisation.csv / E_S7_colocalisation.md).

---

**Supplementary Table S1.** Complete Mendelian randomization results across all exposure–outcome pairs and methods. Inverse-variance weighted (IVW) MR is the primary estimator. MR-Egger, weighted median (WMedian), and weighted mode (WMode) are shown as sensitivity estimators. OR, odds ratio; CI, confidence interval; nSNP, number of harmonised instruments retained after palindromic-variant removal and F > 10 filtering. Cross-cohort pairs use FinnGen R13 exposures and independent GWAS Catalog outcomes; the abdominal aortic aneurysm (AAA) analysis is a prespecified sensitivity check. Full machine-readable tables: E_results_cross_cohort.csv, E_results_cross_cohort_methods.csv, E_results_AAA.csv. S1a. Primary (IVW) and headline sensitivity estimates per pair

| Exposure | Outcome | nSNP | min F | IVW OR (95% CI) | IVW P | Egger OR | Egger int. P | WMedian OR | WMode OR | Q P | Bonferroni P |
|---|---|---|---|---|---|---|---|---|---|---|---|
| ASD | AF | 78 | 30.1 | 1.0145 (1.0006–1.0285) | 0.04066 | 0.9174 | 0.005956 | 0.9959 | 0.995 | 1.06e-31 | 0.4879 |
| ASD | HF | 55 | 30.1 | 1.0002 (0.989–1.0115) | 0.9737 | 0.7038 | 1.86e-05 | 0.9877 | 0.9885 | 0.003386 | 1 |
| ASD | TAA | 61 | 30.1 | 1.0225 (0.9756–1.0717) | 0.3529 | 1.1282 | 0.3965 | 0.9781 | 0.9741 | 0.9998 | 1 |
| BAV | AF | 14 | 33.1 | 1.0464 (1.0311–1.062) | 1.74e-09 | 1.4035 | 0.03148 | 1.0384 | 1.029 | 0.8523 | 2.09e-08 |
| BAV | HF | 10 | 33.1 | 1.0241 (1.0035–1.045) | 0.02155 | 1.2659 | 0.2284 | 1.0235 | 1.0376 | 0.9908 | 0.2586 |
| BAV | TAA | 10 | 33.1 | 0.9354 (0.8406–1.041) | 0.2211 | 1.6825 | 0.5116 | 0.9241 | 0.9732 | 1 | 1 |
| LVOTO | AF | 19 | 29.9 | 1.0648 (1.0423–1.0878) | 8.50e-09 | 1.1769 | 0.3532 | 1.0529 | 1.0442 | 0.7296 | 1.02e-07 |
| LVOTO | HF | 14 | 29.9 | 1.0343 (1.0048–1.0646) | 0.02219 | 1.1205 | 0.5576 | 1.0293 | 1.0201 | 0.9985 | 0.2663 |
| LVOTO | TAA | 14 | 29.9 | 0.8893 (0.7634–1.036) | 0.1322 | 1.5638 | 0.4311 | 0.9069 | 0.9529 | 1 | 1 |
| Overall_CHD | AF | 62 | 30.1 | 1.2949 (1.2572–1.3337) | 5.88e-66 | 1.004 | 4.39e-07 | 1.3092 | 1.3131 | 4.46e-10 | 7.06e-65 |
| Overall_CHD | HF | 50 | 30.1 | 1.1985 (1.1704–1.2273) | 1.72e-50 | 1.4389 | 0.219 | 1.2092 | 1.2098 | 0.9743 | 2.07e-49 |
| Overall_CHD | TAA | 50 | 30.1 | 1.5586 (1.3724–1.7701) | 8.05e-12 | 1.2318 | 0.3139 | 1.7366 | 1.7456 | 1 | 9.66e-11 |
| S1b. All methods, all pairs (48 estimates) |  |  |  |  |  |  |  |  |  |  |  |
| Exposure | Outcome | Method | nSNP | beta | SE | OR (95% CI) | P |  |  |  |  |
| ASD | AF | Inverse variance weighted | 78 | 0.01438 | 0.00702 | 1.0145 (1.0006–1.0285) | 0.04066 |  |  |  |  |
| ASD | AF | MR Egger | 78 | -0.0862 | 0.03618 | 0.9174 (0.8546–0.9848) | 0.01968 |  |  |  |  |
| ASD | AF | Weighted median | 78 | -0.00413 | 0.00473 | 0.9959 (0.9867–1.0052) | 0.3825 |  |  |  |  |
| ASD | AF | Weighted mode | 78 | -0.00496 | 0.00689 | 0.995 (0.9817–1.0086) | 0.4737 |  |  |  |  |
| ASD | HF | Inverse variance weighted | 55 | 0.00019 | 0.00573 | 1.0002 (0.989–1.0115) | 0.9737 |  |  |  |  |
| ASD | HF | MR Egger | 55 | -0.35123 | 0.07485 | 0.7038 (0.6078–0.815) | 1.94e-05 |  |  |  |  |
| ASD | HF | Weighted median | 55 | -0.01235 | 0.00591 | 0.9877 (0.9764–0.9992) | 0.03658 |  |  |  |  |
| ASD | HF | Weighted mode | 55 | -0.01152 | 0.00959 | 0.9885 (0.9701–1.0073) | 0.2348 |  |  |  |  |
| ASD | TAA | Inverse variance weighted | 61 | 0.02227 | 0.02397 | 1.0225 (0.9756–1.0717) | 0.3529 |  |  |  |  |
| ASD | TAA | MR Egger | 61 | 0.1206 | 0.11761 | 1.1282 (0.8959–1.4207) | 0.3093 |  |  |  |  |
| ASD | TAA | Weighted median | 61 | -0.02213 | 0.03102 | 0.9781 (0.9204–1.0394) | 0.4755 |  |  |  |  |
| ASD | TAA | Weighted mode | 61 | -0.02628 | 0.05636 | 0.9741 (0.8722–1.0878) | 0.6427 |  |  |  |  |
| BAV | AF | Inverse variance weighted | 14 | 0.04539 | 0.00754 | 1.0464 (1.0311–1.062) | 1.74e-09 |  |  |  |  |
| BAV | AF | MR Egger | 14 | 0.33897 | 0.12084 | 1.4035 (1.1075–1.7786) | 0.01589 |  |  |  |  |
| BAV | AF | Weighted median | 14 | 0.03771 | 0.01089 | 1.0384 (1.0165–1.0608) | 0.0005352 |  |  |  |  |
| BAV | AF | Weighted mode | 14 | 0.02855 | 0.02 | 1.029 (0.9894–1.0701) | 0.177 |  |  |  |  |
| BAV | HF | Inverse variance weighted | 10 | 0.02378 | 0.01035 | 1.0241 (1.0035–1.045) | 0.02155 |  |  |  |  |
| BAV | HF | MR Egger | 10 | 0.23579 | 0.16288 | 1.2659 (0.9199–1.742) | 0.1858 |  |  |  |  |
| BAV | HF | Weighted median | 10 | 0.02319 | 0.01302 | 1.0235 (0.9977–1.0499) | 0.07501 |  |  |  |  |
| BAV | HF | Weighted mode | 10 | 0.0369 | 0.01909 | 1.0376 (0.9995–1.0771) | 0.08522 |  |  |  |  |
| BAV | TAA | Inverse variance weighted | 10 | -0.06674 | 0.05455 | 0.9354 (0.8406–1.041) | 0.2211 |  |  |  |  |
| BAV | TAA | MR Egger | 10 | 0.52025 | 0.85645 | 1.6825 (0.314–9.015) | 0.5604 |  |  |  |  |
| BAV | TAA | Weighted median | 10 | -0.07897 | 0.06426 | 0.9241 (0.8147–1.0481) | 0.2191 |  |  |  |  |
| BAV | TAA | Weighted mode | 10 | -0.02714 | 0.09785 | 0.9732 (0.8034–1.179) | 0.7878 |  |  |  |  |
| LVOTO | AF | Inverse variance weighted | 19 | 0.06281 | 0.01091 | 1.0648 (1.0423–1.0878) | 8.50e-09 |  |  |  |  |
| LVOTO | AF | MR Egger | 19 | 0.16285 | 0.10537 | 1.1769 (0.9573–1.4468) | 0.1406 |  |  |  |  |
| LVOTO | AF | Weighted median | 19 | 0.05155 | 0.015 | 1.0529 (1.0224–1.0843) | 0.0005915 |  |  |  |  |
| LVOTO | AF | Weighted mode | 19 | 0.04327 | 0.03205 | 1.0442 (0.9806–1.1119) | 0.1938 |  |  |  |  |
| LVOTO | HF | Inverse variance weighted | 14 | 0.03373 | 0.01475 | 1.0343 (1.0048–1.0646) | 0.02219 |  |  |  |  |
| LVOTO | HF | MR Egger | 14 | 0.11375 | 0.13345 | 1.1205 (0.8626–1.4555) | 0.4107 |  |  |  |  |
| LVOTO | HF | Weighted median | 14 | 0.02887 | 0.01903 | 1.0293 (0.9916–1.0684) | 0.1293 |  |  |  |  |
| LVOTO | HF | Weighted mode | 14 | 0.01993 | 0.03043 | 1.0201 (0.9611–1.0828) | 0.524 |  |  |  |  |
| LVOTO | TAA | Inverse variance weighted | 14 | -0.1173 | 0.07791 | 0.8893 (0.7634–1.036) | 0.1322 |  |  |  |  |
| LVOTO | TAA | MR Egger | 14 | 0.44715 | 0.69722 | 1.5638 (0.3988–6.133) | 0.5334 |  |  |  |  |
| LVOTO | TAA | Weighted median | 14 | -0.09778 | 0.09894 | 0.9069 (0.747–1.1009) | 0.3231 |  |  |  |  |
| LVOTO | TAA | Weighted mode | 14 | -0.04826 | 0.15295 | 0.9529 (0.7061–1.286) | 0.7574 |  |  |  |  |
| Overall_CHD | AF | Inverse variance weighted | 62 | 0.25842 | 0.01506 | 1.2949 (1.2572–1.3337) | 5.88e-66 |  |  |  |  |
| Overall_CHD | AF | MR Egger | 62 | 0.00402 | 0.04654 | 1.004 (0.9165–1.0999) | 0.9315 |  |  |  |  |
| Overall_CHD | AF | Weighted median | 62 | 0.2694 | 0.01641 | 1.3092 (1.2677–1.352) | 1.49e-60 |  |  |  |  |
| Overall_CHD | AF | Weighted mode | 62 | 0.27243 | 0.03443 | 1.3131 (1.2275–1.4048) | 6.22e-11 |  |  |  |  |
| Overall_CHD | HF | Inverse variance weighted | 50 | 0.18106 | 0.01212 | 1.1985 (1.1704–1.2273) | 1.72e-50 |  |  |  |  |
| Overall_CHD | HF | MR Egger | 50 | 0.36387 | 0.14729 | 1.4389 (1.0781–1.9204) | 0.0171 |  |  |  |  |
| Overall_CHD | HF | Weighted median | 50 | 0.18994 | 0.01759 | 1.2092 (1.1682–1.2516) | 3.55e-27 |  |  |  |  |
| Overall_CHD | HF | Weighted mode | 50 | 0.19049 | 0.04686 | 1.2098 (1.1037–1.3262) | 0.0001732 |  |  |  |  |
| Overall_CHD | TAA | Inverse variance weighted | 50 | 0.4438 | 0.06491 | 1.5586 (1.3724–1.7701) | 8.05e-12 |  |  |  |  |
| Overall_CHD | TAA | MR Egger | 50 | 0.20845 | 0.24016 | 1.2318 (0.7693–1.9722) | 0.3897 |  |  |  |  |
| Overall_CHD | TAA | Weighted median | 50 | 0.55192 | 0.09111 | 1.7366 (1.4526–2.0761) | 1.38e-09 |  |  |  |  |
| Overall_CHD | TAA | Weighted mode | 50 | 0.55712 | 0.21031 | 1.7456 (1.1559–2.6362) | 0.01083 |  |  |  |  |
| S1c. Abdominal aortic aneurysm sensitivity analysis |  |  |  |  |  |  |  |  |  |  |  |
| Exposure | Outcome | nSNP | min F | IVW OR | IVW P | Egger OR | WMedian OR | WMode OR | Q P |  |  |
| ASD | AAA | 75 | 30.1 | 1.0091 | 0.1507 | 0.9802 | 0.9966 | 0.9954 | 1 |  |  |
| BAV | AAA | 14 | 33.1 | 0.9873 | 0.3694 | 0.8497 | 0.9783 | 0.9742 | 1 |  |  |
| LVOTO | AAA | 19 | 29.9 | 0.9809 | 0.3496 | 0.7575 | 0.97 | 1.0159 | 1 |  |  |
| Overall_CHD | AAA | 58 | 30.1 | 1.0617 | 0.0009098 | 1.2008 | 1.0389 | 1.0348 | 1 |  |  |

**Supplementary Table S2.** MR-PRESSO global heterogeneity and outlier tests. MR-PRESSO (v1.0) was applied with 100 permutations and a significance threshold of 0.05. The Global Test P assesses overall horizontal pleiotropy/heterogeneity; the outlier test reports the number of SNPs flagged. For the primary subtype pairs (BAV→AF, LVOTO→AF) the global test was non-significant (P=0.86 and 0.80) with no outliers, supporting the robustness of the AF findings. For ASD→AF and overall-CHD→AF the global test was significant (P<0.01) although no single outlier was detected, indicating diffuse rather than discrete heterogeneity; this reinforces the decision not to interpret these broad/non-focal exposures as robust findings. AAA sensitivity pairs were not evaluated with the global test in the primary pipeline. Source: E_results_cross_cohort.csv, E_results_AAA.csv.

| Exposure | Outcome | Global test P | No. outliers | Note |
|---|---|---|---|---|
| ASD | AF | <0.01 | 0 | global test significant (<0.01); no discrete outlier |
| ASD | HF | 0.01 | 0 | borderline (P=0.01); no discrete outlier |
| ASD | TAA | 1 | 0 | non-significant; no outliers |
| BAV | AF | 0.86 | 0 | non-significant; no outliers |
| BAV | HF | 1 | 0 | non-significant; no outliers |
| BAV | TAA | 1 | 0 | non-significant; no outliers |
| LVOTO | AF | 0.8 | 0 | non-significant; no outliers |
| LVOTO | HF | 0.99 | 0 | non-significant; no outliers |
| LVOTO | TAA | 1 | 0 | non-significant; no outliers |
| Overall_CHD | AF | <0.01 | 0 | global test significant (<0.01); no discrete outlier |
| Overall_CHD | HF | 1 | 0 | non-significant; no outliers |
| Overall_CHD | TAA | 1 | 0 | non-significant; no outliers |
| AAA sensitivity pairs |  |  |  |  |
| Exposure | Outcome | Global test P | No. outliers | Note |
| ASD | AAA | not evaluated | 0 | not evaluated |
| BAV | AAA | not evaluated | 0 | not evaluated |
| LVOTO | AAA | not evaluated | 0 | not evaluated |
| Overall_CHD | AAA | not evaluated | 0 | not evaluated |

**Supplementary Table S3.**
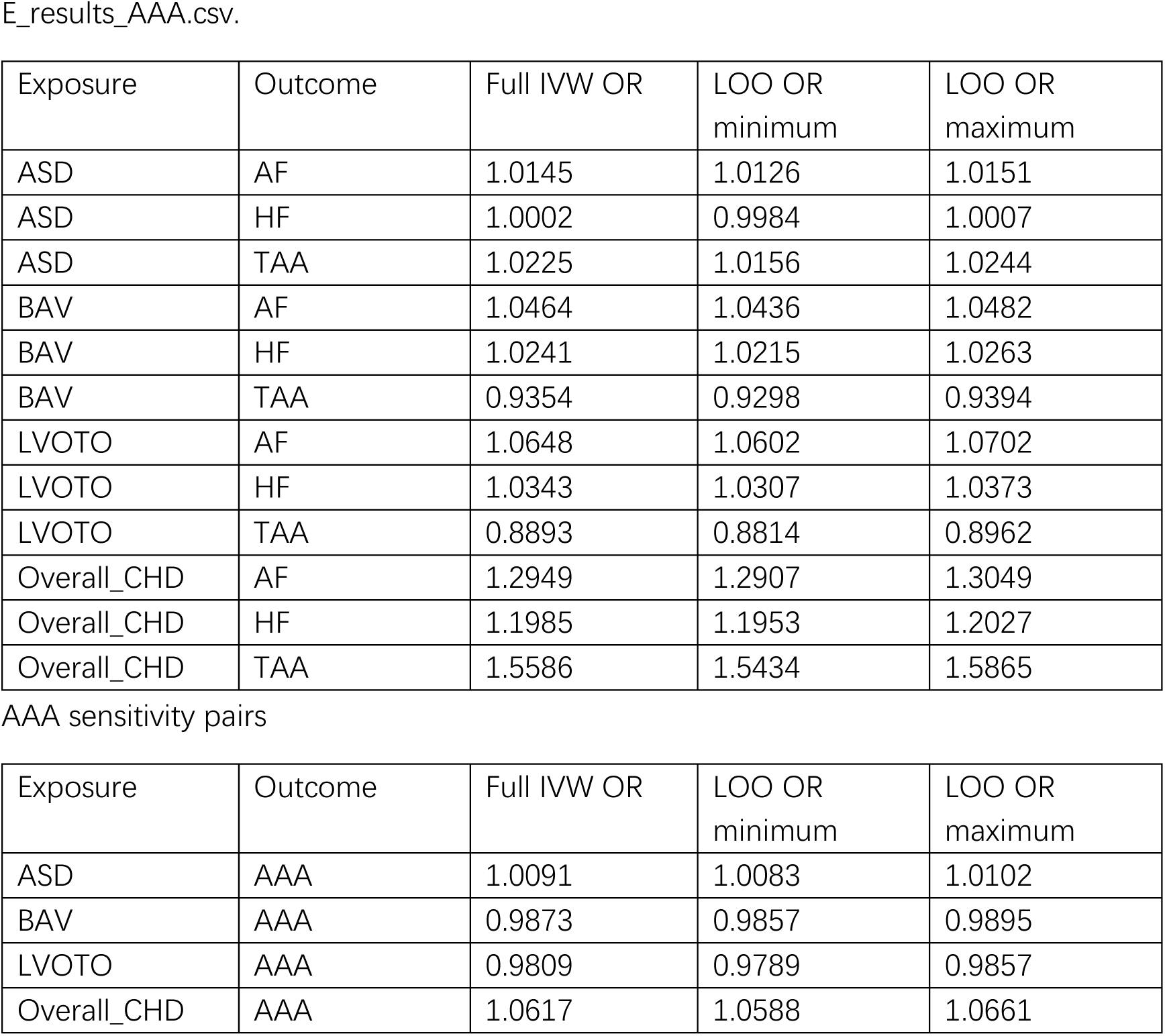
Leave-one-out sensitivity analyses. For each exposure–outcome pair, the IVW odds ratio was recomputed after removing each instrument in turn. The range of resulting ORs (minimum–maximum) is reported; a range that remains close to the full-instrument IVW estimate indicates the result is not driven by a single influential variant. Source: E_results_cross_cohort.csv, E_results_AAA.csv.

| Exposure | Outcome | Full IVW OR | LOO OR minimum | LOO OR maximum |
|---|---|---|---|---|
| ASD | AF | 1.0145 | 1.0126 | 1.0151 |
| ASD | HF | 1.0002 | 0.9984 | 1.0007 |
| ASD | TAA | 1.0225 | 1.0156 | 1.0244 |
| BAV | AF | 1.0464 | 1.0436 | 1.0482 |
| BAV | HF | 1.0241 | 1.0215 | 1.0263 |
| BAV | TAA | 0.9354 | 0.9298 | 0.9394 |
| LVOTO | AF | 1.0648 | 1.0602 | 1.0702 |
| LVOTO | HF | 1.0343 | 1.0307 | 1.0373 |
| LVOTO | TAA | 0.8893 | 0.8814 | 0.8962 |
| Overall_CHD | AF | 1.2949 | 1.2907 | 1.3049 |
| Overall_CHD | HF | 1.1985 | 1.1953 | 1.2027 |
| Overall_CHD | TAA | 1.5586 | 1.5434 | 1.5865 |
| AAA sensitivity pairs |  |  |  |  |
| Exposure | Outcome | Full IVW OR | LOO OR minimum | LOO OR maximum |
| ASD | AAA | 1.0091 | 1.0083 | 1.0102 |
| BAV | AAA | 0.9873 | 0.9857 | 0.9895 |
| LVOTO | AAA | 0.9809 | 0.9789 | 0.9857 |
| Overall_CHD | AAA | 1.0617 | 1.0588 | 1.0661 |

**Supplementary Table S4.** Steiger directionality tests (recomputed) Steiger directionality was recomputed with get_r_from_lor() on the liability scale using endpoint-specific case counts from FinnGen R13 Risteys (BAV 1,356; LVOTO 4,050; ASD 1,443; overall CHD 6,216; total sample 500,186). For binary outcomes the SNP–trait correlation was derived on the liability scale. Correct direction (recomputed) indicates the exposure–outcome orientation is supported. Source: E_results_steiger_fixed.csv.

| Exposure | Outcome | nSNP | r2 exposure | r2 outcome | Steiger P (recomputed) | Correct direction | P (old approximation) | Direction changed |
|---|---|---|---|---|---|---|---|---|
| ASD | AAA | 75 | 0.3118 | 0.0004222 | <1e-300 | TRUE | 2.31e-288 | no |
| ASD | AF | 78 | 0.3089 | 0.003646 | <1e-300 | TRUE | <1e-300 | no |
| ASD | HF | 55 | 0.2376 | 0.0004832 | <1e-300 | TRUE | 4.96e-277 | no |
| ASD | TAA | 61 | 0.2402 | 0.005806 | <1e-300 | TRUE | 4.61e-07 | no |
| BAV | AAA | 14 | 0.05978 | 2.782e-05 | <1e-300 | TRUE | 3.16e-49 | no |
| BAV | AF | 14 | 0.05978 | 0.0001504 | <1e-300 | TRUE | 5.79e-52 | no |
| BAV | HF | 10 | 0.04447 | 3.441e-05 | <1e-300 | TRUE | 7.79e-49 | no |
| BAV | TAA | 10 | 0.04447 | 0.0002682 | 1.08e-162 | TRUE | 0.01245 | no |
| LVOTO | AAA | 19 | 0.02869 | 5.169e-05 | <1e-300 | TRUE | 1.13e-66 | no |
| LVOTO | AF | 19 | 0.02869 | 0.0001611 | <1e-300 | TRUE | 4.31e-76 | no |
| LVOTO | HF | 14 | 0.02191 | 3.789e-05 | <1e-300 | TRUE | 1.53e-70 | no |
| LVOTO | TAA | 14 | 0.02191 | 0.0004205 | 5.68e-70 | TRUE | 0.003896 | no |
| Overall_CHD | AAA | 58 | 0.04435 | 0.0003501 | <1e-300 | TRUE | 3.73e-149 | no |
| Overall_CHD | AF | 62 | 0.04587 | 0.003556 | <1e-300 | TRUE | 4.74e-87 | no |
| Overall_CHD | HF | 50 | 0.0389 | 0.001503 | <1e-300 | TRUE | 4.33e-122 | no |
| Overall_CHD | TAA | 50 | 0.03718 | 0.0123 | 8.77e-31 | TRUE | 0.9287 | YES |

**Supplementary Table S5.** Biological annotation of BAV and LVOTO lead instruments. Nearest gene from FinnGen R13 summary statistics (nearest_genes field, GRCh38), obtained by tabix query of the endpoint summary-statistic files. The great majority of lead instruments map to a single 1p22 locus annotated LINC01708, adjacent to PALMD; several of these SNPs (rs6702619, rs11166276, rs7543130) are reported eQTLs for PALMD in aortic valve tissue and this locus is an established BAV / aortic-root size association signal. This concentration of instruments in one developmental locus is noted explicitly as a limitation of biological breadth. Nearest-gene assignment is indicative and does not by itself establish the causal gene or eQTL target. We therefore performed Bayesian colocalization of this locus with PALMD cis-eQTLs; the result was negative (all PP4 < 0.003), reported in Supplementary Note 1 and Supplementary Table S7. Instrument flow and why the annotated set differs from the analysis set. FinnGen R13 yielded 16 genome-wide significant instruments for BAV and 22 for LVOTO (Table 1). Of these, one per subtype at chr1:99,567,756 lacked an rsID and could not be matched to the GWAS Catalog outcomes, leaving 15 BAV and 21 LVOTO rsID-bearing instruments annotated in this table. For the primary AF analysis, one LVOTO instrument (rs1473251324, an indel at 17:78,242,643 with beta=1.542; minor allele frequency 0.06%) was not present in the AF outcome GWAS and therefore did not enter the analysis; a further instrument per subtype was removed by palindromic-SNP harmonisation (TwoSampleMR action=2, effect-allele frequency 0.42–0.58), leaving the final counts of 14 (BAV) and 19 (LVOTO) used for the AF estimates in Table 2. Thus the numbers are BAV 16→15→14 and LVOTO 22→21→19 at the annotation, outcome-matching, and harmonisation stages, respectively. Note that rs1473251324 was excluded by outcome matching and, independently, falls below the 1% minor-allele-frequency threshold, so its extreme effect does not influence any reported AF estimate.

| Subtype | SNP | Chr | Pos (hg38) | EA/OA | beta | P | Nearest gene | Locus / annotation |
|---|---|---|---|---|---|---|---|---|
| BAV | rs4581299 | 1 | 99570504 | T/A | 0.236236 | 6.35521e-09 | LINC01708 | 1p22 LINC01708/PALMD |
| BAV | rs7554464 | 1 | 99570980 | C/G | 0.236839 | 5.8039e-09 | LINC01708 | 1p22 LINC01708/PALMD |
| BAV | rs1890753 | 1 | 99573963 | C/G | 0.249812 | 4.72825e-11 | LINC01708 | 1p22 LINC01708/PALMD |
| BAV | rs9660000 | 1 | 99576082 | A/G | 0.236713 | 5.76355e-09 | LINC01708 | 1p22 LINC01708/PALMD |
| BAV | rs11166276 | 1 | 99579683 | T/C | 0.266299 | 2.19028e-12 | LINC01708 | 1p22 LINC01708/PALMD; PALMD eQTL (aortic valve) |
| BAV | rs6702619 | 1 | 99580690 | G/T | 0.264928 | 2.83335e-12 | LINC01708 | 1p22 LINC01708/PALMD; PALMD eQTL (aortic valve) |
| BAV | rs12025235 | 1 | 99582777 | T/C | 0.237777 | 4.89846e-09 | LINC01708 | 1p22 LINC01708/PALMD |
| BAV | rs7543039 | 1 | 99584092 | T/C | 0.265458 | 2.55094e-12 | LINC01708 | 1p22 LINC01708/PALMD |
| BAV | rs7543130 | 1 | 99584229 | A/C | 0.265471 | 2.54214e-12 | LINC01708 | 1p22 LINC01708/PALMD; PALMD eQTL (aortic valve) |
| BAV | rs2893418 | 1 | 99586112 | A/G | 0.237785 | 4.88754e-09 | LINC01708 | 1p22 LINC01708/PALMD |
| BAV | rs1856616 | 1 | 99586424 | T/C | 0.23134 | 3.95112e-09 | LINC01708 | 1p22 LINC01708/PALMD |
| BAV | rs6693824 | 1 | 99588041 | C/G | 0.227847 | 6.63835e-09 | LINC01708 | 1p22 LINC01708/PALMD |
| BAV | rs954906 | 1 | 99588425 | T/C | 0.227847 | 6.63835e-09 | LINC01708 | 1p22 LINC01708/PALMD |
| BAV | rs1339860 | 1 | 99589243 | T/A | 0.227847 | 6.63835e-09 | LINC01708 | 1p22 LINC01708/PALMD |
| BAV | rs2392041 | 1 | 99590669 | A/G | 0.233663 | 8.75629e-09 | LINC01708 | 1p22 LINC01708/PALMD |
| LVOTO | rs2027357 | 1 | 99566971 | G/C | 0.13747 | 3.67012e-08 | LINC01708 | 1p22 LINC01708/PALMD |
| LVOTO | rs7513849 | 1 | 99567280 | A/G | 0.137628 | 3.97667e-08 | LINC01708 | 1p22 LINC01708/PALMD |
| LVOTO | rs4581299 | 1 | 99570504 | T/A | 0.145041 | 8.64072e-10 | LINC01708 | 1p22 LINC01708/PALMD |
| LVOTO | rs7554464 | 1 | 99570980 | C/G | 0.144534 | 9.84124e-10 | LINC01708 | 1p22 LINC01708/PALMD |
| LVOTO | rs1890753 | 1 | 99573963 | C/G | 0.16621 | 8.30615e-14 | LINC01708 | 1p22 LINC01708/PALMD |
| LVOTO | rs9660000 | 1 | 99576082 | A/G | 0.144472 | 9.74406e-10 | LINC01708 | 1p22 LINC01708/PALMD |
| LVOTO | rs11166276 | 1 | 99579683 | T/C | 0.168252 | 4.23156e-14 | LINC01708 | 1p22 LINC01708/PALMD; PALMD eQTL (aortic valve) |
| LVOTO | rs6702619 | 1 | 99580690 | G/T | 0.16844 | 3.91201e-14 | LINC01708 | 1p22 LINC01708/PALMD; PALMD eQTL (aortic valve) |
| LVOTO | rs12025235 | 1 | 99582777 | T/C | 0.144211 | 1.03569e-09 | LINC01708 | 1p22 LINC01708/PALMD |
| LVOTO | rs7543039 | 1 | 99584092 | T/C | 0.168319 | 4.08131e-14 | LINC01708 | 1p22 LINC01708/PALMD |
| LVOTO | rs7543130 | 1 | 99584229 | A/C | 0.168171 | 4.28746e-14 | LINC01708 | 1p22 LINC01708/PALMD; PALMD eQTL (aortic valve) |
| LVOTO | rs2893418 | 1 | 99586112 | A/G | 0.144032 | 1.08253e-09 | LINC01708 | 1p22 LINC01708/PALMD |
| LVOTO | rs1856616 | 1 | 99586424 | T/C | 0.139297 | 1.1908e-09 | LINC01708 | 1p22 LINC01708/PALMD |
| LVOTO | rs6693824 | 1 | 99588041 | C/G | 0.135435 | 3.31352e-09 | LINC01708 | 1p22 LINC01708/PALMD |
| LVOTO | rs954906 | 1 | 99588425 | T/C | 0.135435 | 3.31352e-09 | LINC01708 | 1p22 LINC01708/PALMD |
| LVOTO | rs1339860 | 1 | 99589243 | T/A | 0.135435 | 3.31352e-09 | LINC01708 | 1p22 LINC01708/PALMD |
| LVOTO | rs2392041 | 1 | 99590669 | A/G | 0.140117 | 2.95189e-09 | LINC01708 | 1p22 LINC01708/PALMD |
| LVOTO | rs946706 | 1 | 99596278 | A/G | 0.133857 | 4.60013e-08 | LINC01708 | 1p22 LINC01708/PALMD |
| LVOTO | rs6672117 | 1 | 99596861 | G/A | 0.133884 | 4.5732e-08 | LINC01708 | 1p22 LINC01708/PALMD |
| LVOTO | rs28460841 | 1 | 241948609 | A/G | -0.203576 | 1.48109e-08 | NA | unannotated |
| LVOTO | rs1473251324 | 17 | 78242643 | ACAG/A | 1.54163 | 4.72531e-08 | TMEM235 | TMEM235 |

Summary: 34 of the 36 rsID-bearing instruments listed here map to the 1p22 LINC01708/PALMD locus (BAV 15/15; LVOTO 19/21); the two remaining LVOTO instruments are unannotated (rs28460841) or map to TMEM235 (rs1473251324, excluded from AF analysis as described above). The table lists 36 of the 38 FinnGen genome-wide significant instruments; the two omitted variants lacked rsIDs. Final harmonised analysis counts were 14 (BAV) and 19 (LVOTO).

**Supplementary Table S6.** Supplementary Table S6. Methods note — reconstruction of standard errors for the abdominal aortic aneurysm GWAS.

Purpose. The harmonised abdominal aortic aneurysm (AAA) GWAS (GWAS Catalog GCST90475994; 16,289 cases / 427,111 controls) was analysed as a prespecified sensitivity check of the phenotype-mismatch explanation for aortopathy non-replication. Its harmonised release reported no standard_error field (all values missing), so standard errors for the log odds ratio were recovered from the reported 95% confidence intervals.

Method. For each instrument, with lower and upper bounds of the 95% CI denoted CI_lower and CI_upper:/n

- beta = ln(OR)

- SE[log(OR)] = [ln(CI_upper) − ln(CI_lower)] / 3.92

The divisor 3.92 approximates 2 × 1.96, the width of a 95% confidence interval on the log scale under a normal approximation. This is a standard, deterministic reconstruction when only point estimates and 95% CIs are available.

Scope. These reconstructed standard errors were used only for the AAA sensitivity analysis (Table S1c, Table S2/S3 AAA panels). They did not affect the primary AF results, which used GWAS Catalog harmonised estimates with reported standard errors directly. All primary estimates and conclusions are unchanged by this reconstruction.

**Supplementary Table S7.** Bayesian colocalisation of the 1p22 locus with PALMD cis-eQTLs. coloc.abf (v5.2.3; p1=p2=1e-4, p12=1e-5) between each CHD-subtype GWAS (FinnGen R13, chr1:99.0-100.2 Mb, GRCh38) and PALMD (ENSG00000099260) cis-eQTLs from three GTEx tissues in the eQTL Catalogue. Effect alleles harmonised; ambiguous/palindromic variants removed. PP4 >= 0.8 supports a shared causal variant. All six pairs have PP4 < 0.003: no support for PALMD as the effector gene; posterior mass lies on H3 (two distinct variants) or H1 (GWAS signal without a matched eQTL).

| Pair | nSNP | PP.H0 | PP.H1 | PP.H2 | PP.H3 | PP.H4 | PP4/PP3 | Verdict |
|---|---|---|---|---|---|---|---|---|
| BAV / aorta | 3869 | 4.054e-06 | 0.9742 | 9.567e-08 | 0.02299 | 0.002786 | 0.121 | No support (PP4<0.8) |
| BAV / atrium | 3859 | 3.879e-08 | 0.009323 | 4.122e-06 | 0.9906 | 4.051e-05 | 4.09e-05 | No support (PP4<0.8) |
| BAV / lv | 3853 | 2.695e-20 | 6.477e-15 | 4.161e-06 | 1 | 2.615e-08 | 2.62e-08 | No support (PP4<0.8) |
| LVOTO / aorta | 3869 | 6.474e-08 | 0.9743 | 1.528e-09 | 0.02299 | 0.002749 | 0.12 | No support (PP4<0.8) |
| LVOTO / atrium | 3859 | 6.196e-10 | 0.009324 | 6.583e-08 | 0.9906 | 3.998e-05 | 4.04e-05 | No support (PP4<0.8) |
| LVOTO / lv | 3853 | 4.304e-22 | 6.477e-15 | 6.646e-08 | 1 | 2.804e-10 | 2.8e-10 | No support (PP4<0.8) |

## References

[1] : Hoffman JI, Kaplan S. The incidence of congenital heart disease. J Am Coll Cardiol. 2002;39(12):1890–1900. doi:10.1016/s0735-1097(02)01886-7.

[2] : Marelli AJ, Ionescu-Ittu R, Mackie AS, Guo L, Dendukuri N, Kaouache M. Lifetime prevalence of congenital heart disease in the general population from 2000 to 2010. Circulation. 2014;130(9):749–756. doi:10.1161/CIRCULATIONAHA.113.008396.

[3] : Baumgartner H, De Backer J, Babu-Narayan SV, et al. 2020 ESC Guidelines for the management of adult congenital heart disease. Eur Heart J. 2021;42(6):563–645. doi:10.1093/eurheartj/ehaa554.

[4] : Bouchardy J, Chatterjee A, Corsello A, et al. Atrial fibrillation in adults with congenital heart disease: a study based on the International CONGENITAL HEART DISEASE (CHD) Registry. Eur Heart J Cardiovasc Imaging. 2021;22(5):560–568. doi:10.1093/ehjci/jeaa327.

[5] : Davies HA, Bowness JS, Brown IA, et al. Management of bicuspid aortic valve disease: consensus document from the British Heart Valve Society and British Association for Cardiovascular Prevention and Rehabilitation. Open Heart. 2021;8(2):e001800. doi:10.1136/openhrt-2021-001800.

[6] : Forbes TJ, Kim DW, Du W, et al. Comparison of surgical, stent, and balloon angioplasty treatment of native coarctation of the aorta: an observational study by the CCISC (Congenital Cardiovascular Interventional Study Consortium). J Am Coll Cardiol. 2011;58(26):2664–2674. doi:10.1016/j.jacc.2011.08.053.

[7] : Lawlor DA, Harbord RM, Sterne JAC, Timpson N, Davey Smith G. Mendelian randomization: using genes as instruments for making causal inferences in epidemiology. Stat Med. 2008;27(8):1133–1163. doi:10.1002/sim.3034.

[8] : Jing X, Cao Y, Wang Q. Exploring the causal association between congenital heart disease and stroke based on two-sample Mendelian randomization. Cardiovasc Diagn Ther. 2025;15(1):61–77. doi:10.21037/cdt-24-422. PMID: 40115093. (Reported congenital anomalies of great vessels → AF OR 1.07, 95% CI 1.02–1.12.)

[9] : Skrivankova VW, Richmond RC, Woolf BAR, et al. Strengthening the reporting of observational studies in epidemiology using mendelian randomisation (STROBE-MR): explanation and elaboration. BMJ. 2021;375:n2233. doi:10.1136/bmj.n2233.

[10] : Staiger DO, Stock JH. Instrumental variables regression with weak instruments. Econometrica. 1997;65(3):557–586. doi:10.2307/2171753.

[11] : Burgess S, Butterworth A, Thompson SG. Mendelian randomization analysis with multiple genetic variants using summarized data. Genet Epidemiol. 2013;37(7):658–665. doi:10.1002/gepi.21758.

[12] : Bowden J, Davey Smith G, Burgess S. Mendelian randomization with invalid instruments: effect estimation and bias detection through Egger regression. Int J Epidemiol. 2015;44(2):512–525. doi:10.1093/ije/dyv080. Bowden J, Davey Smith G, Haycock PC, Burgess S. Consistent estimation in Mendelian randomization with some invalid instruments using a weighted median estimator. Genet Epidemiol. 2016;40(4):304–314. doi:10.1002/gepi.21965.

[13] : Hartwig FP, Davey Smith G, Bowden J. Robust inference in summary data Mendelian randomization via the zero modal pleiotropy assumption. Int J Epidemiol. 2017;46(6):1985–1998. doi:10.1093/ije/dyx102.

[14] : Verbanck M, Chen CY, Neale B, Do R. Detection of widespread horizontal pleiotropy in causal relationships inferred from Mendelian randomization between complex traits and diseases. Nat Genet. 2018;50(5):693–698. doi:10.1038/s41588-018-0099-7.

[15] : Hemani G, Tilling K, Davey Smith G. Orienting the causal relationship between imprecisely measured traits using GWAS summary data. PLoS Genet. 2017;13(11):e1007081. doi:10.1371/journal.pgen.1007081.

[16] : Burgess S, Davies NM, Thompson SG. Bias due to participant overlap in two-sample Mendelian randomization. Genet Epidemiol. 2016;40(7):597–608. doi:10.1002/gepi.21998.

[17] : Sanderson E, Glymour MM, Holmes MV, et al. Mendelian randomization. Nat Rev Methods Primers. 2022;2:6. doi:10.1038/s43586-021-00092-5.

[18] : FinnGen endpoint definition repository (Risteys), endpoint I9_AORTVALV_EARLY “Aortic valve malfunction, age under 60 years” (precondition EVENT_AGE<60; first used in datafreeze DF9). https://r13.risteys.finngen.fi/endpoint/I9_AORTVALV_EARLY (accessed 2026-11-17).

[19] : Tcheandjieu C, Zanetti D, Yu M, Priest JR. Inherited extremes of aortic diameter confer risk for a specific class of congenital heart disease. Circ Genom Precis Med. 2020;13(6):e003170. doi:10.1161/CIRCGEN.120.003170. PMID: 33191768.

